# Father absence and trajectories of offspring mental health across adolescence and young adulthood: findings from a UK-birth cohort

**DOI:** 10.1101/2021.08.25.21262549

**Authors:** Iryna Culpin, Hein Heuvelman, Dheeraj Rai, Rebecca M Pearson, Carol Joinson, Jon Heron, Jonathan Evans, Alex S F Kwong

**Affiliations:** Centre for Academic Mental Health, Population Health Sciences, Bristol Medical School, University of Bristol, Bristol, United Kingdom; Leeds Institute of Health Sciences, School of Medicine, University of Leeds, United Kingdom; NIHR Biomedical Research Centre, University of Bristol, Bristol, United Kingdom; Avon and Wiltshire Partnership National Health Service (NHS) Trust, Bristol, UK; Centre for Academic Child Health, Population Health Sciences, Bristol Medical School, University of Bristol, Bristol, United Kingdom; MRC Integrative Epidemiology Unit, University of Bristol, Bristol, United Kingdom; Division of Psychiatry, Centre for Clinical Brain Sciences, University of Edinburgh, Edinburgh, United Kingdom

**Keywords:** ALSPAC, biological father absence, offspring depression, trajectories of depressive symptoms, population-based study

## Abstract

**Background:** High prevalence of parental separation and resulting biological father absence raises important questions regarding its impact on offspring mental health across the life course. However, few studies have examined prospective associations between biological father absence in childhood and risk of offspring depression and depressive symptoms trajectories across adolescence and young adulthood. We specifically examined whether these relationships vary by sex and the timing of exposure to father absence (early or middle childhood).

**Methods:** This study is based on up to 8,409 children from the Avon Longitudinal Study of Parents and Children (ALSPAC). Participants provided self-reports of depression (Clinical Interview Schedule-Revised) at age 24 years and depressive symptoms (Short Mood and Feelings Questionnaire) between the ages of 10 and 24 years. Biological father absence in childhood was assessed through maternal questionnaires at regular intervals from birth to 10 years. We used logistic regression to examine the association between biological father absence and depression/depressive symptoms at age 24. We estimated the association between biological father absence and trajectories of depressive symptoms using multilevel growth-curve modelling.

**Results:** Early but not middle childhood father absence was strongly associated with increased odds of offspring depression and greater depressive symptoms at age 24 years. Early childhood father absence was associated with higher trajectories of depressive symptoms during adolescence and early adulthood compared with father presence. Differences in the level of depressive symptoms between middle childhood father absent and father present groups narrowed into early adulthood. Girls whose father was absent in early childhood, compared with present, manifested higher levels of depressive symptoms throughout adolescence, but this difference narrowed by early adulthood. In contrast, boys who experienced father absence in early childhood had similar trajectories of depressive symptoms compared to the father present group but experienced a steep increase in early adulthood. Girls whose fathers were absent in middle childhood manifested higher trajectories across middle adolescence into young adulthood compared to the father present group.

**Conclusions:** We found evidence that father absence in childhood is persistently associated with offspring depression in adolescence and early adulthood and that this relationship varies by sex and timing of father’s departure. Further research is needed to examine whether this relationship is causal and to identify mechanisms that could inform preventative interventions to reduce the risk of depression in children who experience father absence.

## Introduction

Recent evidence suggests that a substantial proportion of marriages (42%)^1^ and cohabiting relationships (27%)^2^ in the UK are likely to end in divorce and separation, with around half of these divorces and separations expected to occur in the first 10 years. It is estimated that the majority of children reside with the mother (74.3% in UK) following divorce and/or parental separation.^1,3^ The high prevalence of parental divorce/separation and the ensuing absence of the biological father from the household raises important questions regarding its impact on offspring mental health at different developmental time points and across the life course. Ample longitudinal evidence emphasises the adverse impact of parental divorce, separation and biological father absence on child and adolescent mental health, whilst accounting for sociodemographic and familial factors.^4-7^ However, it remains unclear whether father absence during childhood is associated with increased risk of mental health difficulties in adulthood.^5^

The association between biological father absence and mental health may depend on the child’s sex and developmental stage when the father left.^8,9^ It has been suggested that father absence that occur during early childhood (birth to 5 years) is associated with higher risk of depression in adolescence than paternal absence during middle childhood (5-10 years).^7,10,11^ However, these findings are inconsistent, and, importantly, it remains unknown whether adverse effects of father absence timing persist into early adulthood.^6^ The evidence with regard to sex differences in the association between father absence and offspring depression is similarly inconclusive,^6^ with some studies reporting stronger effects for females,^7,8^ whilst other studies suggest that males are worse affected.^12,13^

Offspring mental health is a dynamic developmental process that unfolds between individuals and their context.^14^ Thus, it is important to assess individual variation in changes in mental health over time following childhood father absence. Explicit longitudinal modelling of mental health trajectories can assess individual variation across development and estimate changes in mental health over time across childhood into adulthood.^15^ In addition, it enables estimation of age-specific effects (early-*versus* middle-childhood), their persistence effects over time, and interaction with other characteristics such as sex.^16^

Several studies have examined trajectories of offspring mental health following parental separation. Ge et al.^16^ found that children from divorced families had higher trajectories of depressive symptoms across adolescence compared to children from nondivorced families, and that this effect was stronger for females. Cherlin et al.^17^ found a pronounced adverse effect of parental divorce in adolescence on offspring trajectories of depressive symptoms between ages 7 and 33 years suggesting long lasting effects well into adulthood. Similarly, Costello et al.^18^ reported that children from separated families were at higher risk of belonging to a higher trajectory of depressive symptoms from ages 12 to 25 years. However, these findings are limited by the selective and relatively small sample size,^16^ use of depression measures not validated in adolescence,^18^ and relatively large gaps between assessment.^17^ Crucially, none of the studies have explicitly examined the association between biological father absence and depressive symptom trajectories, nor have they looked at the timing of father absence, or sex differences in this association.

In the current study we address this gap in literature using data from a large UK-based birth cohort, the Avon Longitudinal Study of Parents and Children (ALSPAC), to estimate the association between biological father absence in childhood and population trajectories of depressive symptoms between ages 10 and 24 years using multilevel growth-curve modelling. We used longitudinal data with frequent (by-yearly) assessments of depressive symptoms throughout early childhood, adolescence and early adulthood using well-validated measures of child and adolescent psychopathology and adjusting for a range of confounding factors preceding father departure, including marital conflict.

Our specific research questions were:

1. Are offspring whose biological fathers were absent in early (birth to 5 years) and middle (5-10 years) childhood at increased risk of depression at age 24 years, and does this risk vary by sex and timing of father absence?
2. Do trajectories of depressive symptoms between the ages 10 and 24 years for children whose biological fathers were present or absent in early (birth to 5 years) and middle (5-10 years) childhood differ?

## Methods

### Study cohort

The sample comprised participants from the Avon Longitudinal Study of Parents and Children (ALSPAC). During Phase I enrolment, 14,541 pregnant mothers residing in the former Avon Health Authority in the South-West of England with expected dates of delivery between 1 April 1991 and 31 December 1992 were recruited. The total enrolled ALSPAC sample size is 15,454 pregnancies, of which 14,901 were alive at 1 year of age. Ethical approval for the study was obtained from the ALSPAC Ethics and Law Committee and the Local Research Ethics Committees. The study website (www.bristol.ac.uk/alspac/) contains details of all the data that is available through a fully searchable data dictionary and variable search tool: http://www.bris.ac.uk/alspac/researchers/our-data. Further details on the cohort profile, representativeness and phases of recruitment are described in three cohort-profile papers.^19-21^ Study data gathered from participants at 22 years and onwards were collected and managed using REDCap electronic data capture tools hosted at the University of Bristol.^22^

### Measures

#### Exposure: biological father absence in childhood

Absence of the biological father was assessed through maternal self-reported questionnaires at regular intervals since the birth of the study child (full details in Methods S1, Supplementary). The questions enquired whether the present live-in father figure is the natural father of the study child, and if not, how old the study child was when the biological father stopped living with the family. Data on biological father absence were divided into two distinct age periods to capture father absence during early (birth-5 years) and middle (5-10 years) childhood.

#### Outcome: offspring depression and depressive symptoms

Offspring depression was assessed using the computerised version of the Clinical Interview Schedule-Revised (CIS-R),^23^ a fully structured psychiatric interview widely used in the community samples.^24^ It was administered at the research clinic at age 24 years to identify individuals with an ICD-10 diagnosis of depression (*versus* no diagnosis).

Offspring depressive symptoms were assessed using the Short Mood and Feelings Questionnaire (SMFQ)^25^ on nine occasions between ages 10 and 24 years (full details in Methods S1, Supplementary). Scores for the individual items were summed up to produce a summary score (range 0-26). A binary variable was also derived with a cut-off point of ≥10, shown to have high sensitivity and specificity for diagnosed depression,^26^ to identify individuals with depressive symptoms at age 24 years (*versus* no depressive symptoms). The SMFQ has been validated in late adolescence^27^ and correlates highly with the Diagnostic Interview Schedule for Children.^28^ Details on the use of SMFQ in longitudinal models have been reported previously.^29^

### Potential confounders

Disadvantaged socioeconomic status and marital conflict are strong risk factors for biological father absence and offspring depression.^6,9^ Analyses were adjusted for a range of potential confounding factors collected prospectively from maternal antenatal questionnaires, including: financial problems (occurrence of major financial problems since pregnancy *versus* none); maternal educational attainment (three-level scale: minimal education or none, compulsory secondary level (up to age 16 years; O-Level), A Levels/university degree), parental occupational class (professional/managerial *versus* manual), homeownership status (three-level scale: owned/mortgaged, privately-rented, council-rented), maternal depression (18 weeks gestation) assessed using the Edinburgh Postnatal Depression Scale (EPDS; sum-score),^30^ and a measure of parental conflict with higher scores indicating higher levels of inter-parental conflict.

### Statistical analyses

Full details of the statistical analyses, including trajectories, are presented in Methods S1, Supplementary. In summary, we examined characteristics of the sample (Table S1) and prevalence of depression diagnosis and depressive symptoms by the presence or absence of the biological father (Table S2, Supplementary). We used binary logistic regression (*logit* command) to examine the association between father absence occurring during different periods in childhood (early: birth-5 and middle: 5-10 years) and ICD-10 depression diagnosis and depressive symptoms at age 24 years. First, we compared children exposed to father absence from birth-5 years (n=1,943) with all children whose fathers were present between birth-10 years (n=9,003). We included in this group the 726 children whose fathers were absent during the period 5–10 years because, by definition, they were not absent during the first 5 years of life. Second, we compared children exposed to father absence during the period 5–10 years (n=726) with those whose fathers were present throughout the period 0–10 years (n=8,277). We then tested for an interaction between father absence and sex using the likelihood ratio test in the full sample to maximise power to detect interaction effect. We estimated models unadjusted and adjusted for the confounding factors.

Finally, we examined trajectories of depressive symptoms (continuous SMFQ scores) between the ages of 10 and 24 years among those whose fathers were absent/present and then again separately for males and females using multilevel growth-curve models with random intercepts and random slopes to examine how depressive symptoms changed across development for each father absence model (details in Methods S1, Supplementary). We ran both adjusted and unadjusted models to match the depression diagnosis analyses. We included linear, quadratic, cubic and quartic polynomial age terms in the models to accommodate the non-linearity of the trajectories as per previous research.^31^ We interacted the age terms with the main effect of early and middle childhood father absence in two separate models, followed by age terms interacted with sex across the two separate models to create trajectories for each risk group (Results S1, Supplementary). To aid interpretation of the non-linear growth curves, we calculated the predicted mean scores for each trajectory at ages 12, 16, 20 and 24 years, followed by calculating the predicted mean difference at the varying ages between trajectories. To correct for multiple comparison testing with the predicted mean difference analysis, p-values were corrected using the false discovery rate. All analyses were conducted using Stata v.15/MP (StataCorp., USA),^32^ including trajectories estimated using the *runmlwin* command.^33^

### Missing data: multiple imputation, full information maximum likelihood and inverse probability weighted analysis

Characteristics of the study sample by completeness of the data are presented in Results S1 and Table S3, Supplementary. We conducted sensitivity analyses to examine the impact of missing data on our findings. Full description of the imputation methods, including trajectories (full information maximum likelihood and inverse probability analysis to handle missing data), are presented in Method S1, Supplementary.

## Results

### Study sample derivation

Figure 1 represents sample sizes with complete data on exposure, outcome and confounding factors to examine the effects of biological father absence on offspring depression diagnosis and depressive symptoms at age 24 years (full details in Results S1, Supplementary). The sample sizes for those with data on exposure, confounding factors and at least one measurement of depressive symptoms to establish trajectories was 6,020 (father absence birth-5 years) and 5,352 (father absence 5-10 years). The majority of fathers were absent during the first five years of child’s life (17,7%), with the proportion of absent fathers declining between ages 5-10 years (6.6%).

**Figure 1.**
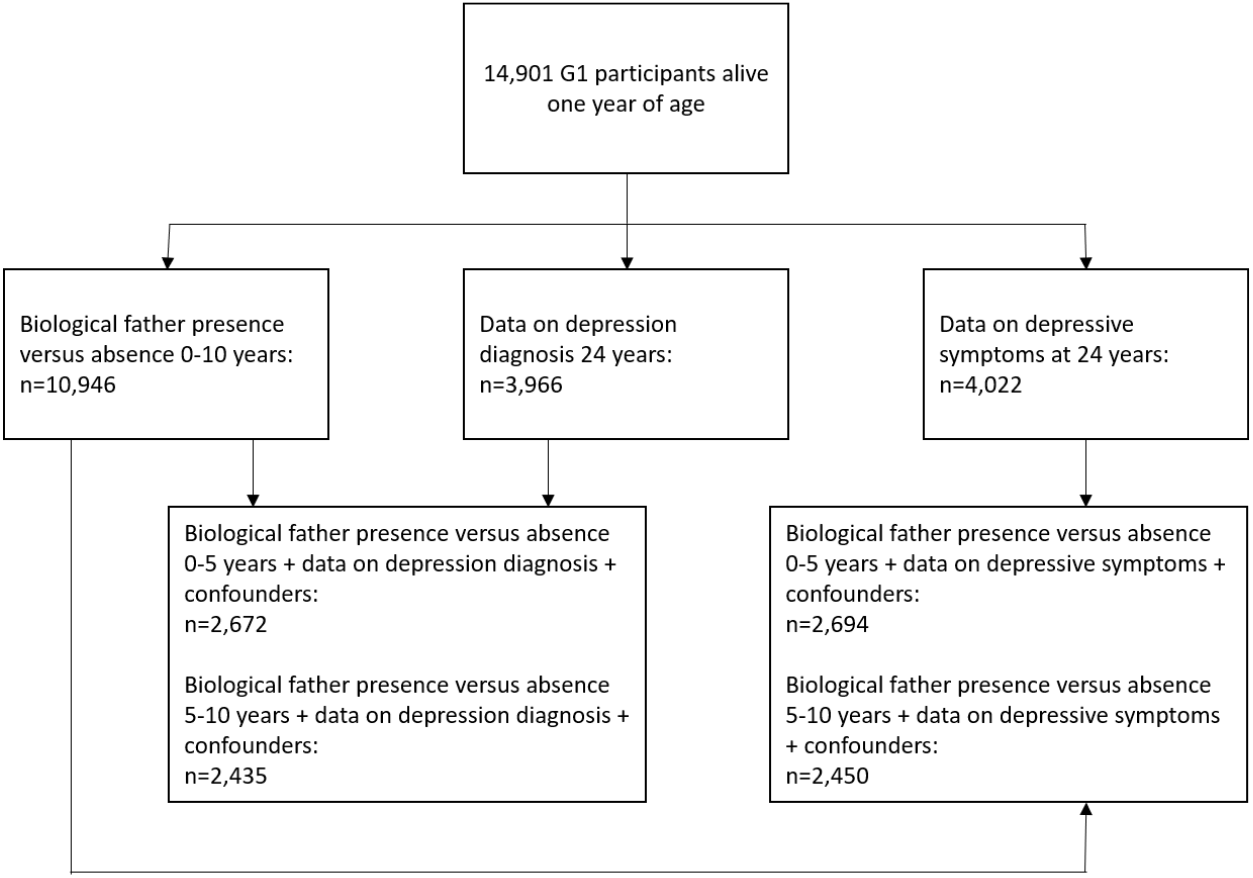
Study sample

### Association between father absence in childhood and depression (diagnosis and symptoms) at age 24 years

Distribution of depression, socioeconomic and familial characteristics in father-present and father-absent samples is presented in Results S1 and Tables S1-S2. Of 3,966 young adults with data on depression diagnosis and 4,022 with data on depressive symptoms at age 24 years, 432 (10.9%, 95%CI 0.09, 0.12) met the criteria for depression, and 994 (24.7%, 95%CI 0.23, 0.26) reported depressive mood. First, we estimated the main effect of father absence and the interaction between father absence and sex in the full sample to maximise power to detect possible interactions. There was evidence of the main effect of father absence in early (OR: 1.97, 95%CI 1.49, 2.60, p≤0.001; Table 1), but not middle childhood (OR: 1.13, 95%CI 0.73, 1.76, p=0.585; Table 1), on depression diagnosis at age 24 years. There was no evidence for an interaction between sex and father absence in early childhood, indicating that the association between early childhood father absence and diagnosis of depression at age 24 years is similar for boys and girls (Table 1). There was evidence for a main effect of father absence in early childhood (OR: 1.54, 95%CI 1.21, 1.93, p≤0.001) and middle childhood (OR: 1.43, 95% CI 1.10, 1.95, p=0.020) on depressive symptoms at age 24 years (Table 1). There was weak evidence for an interaction between sex and father absence in middle, but not early childhood, on depressive symptoms at age 24 years (Table 1).

**Table 1.**
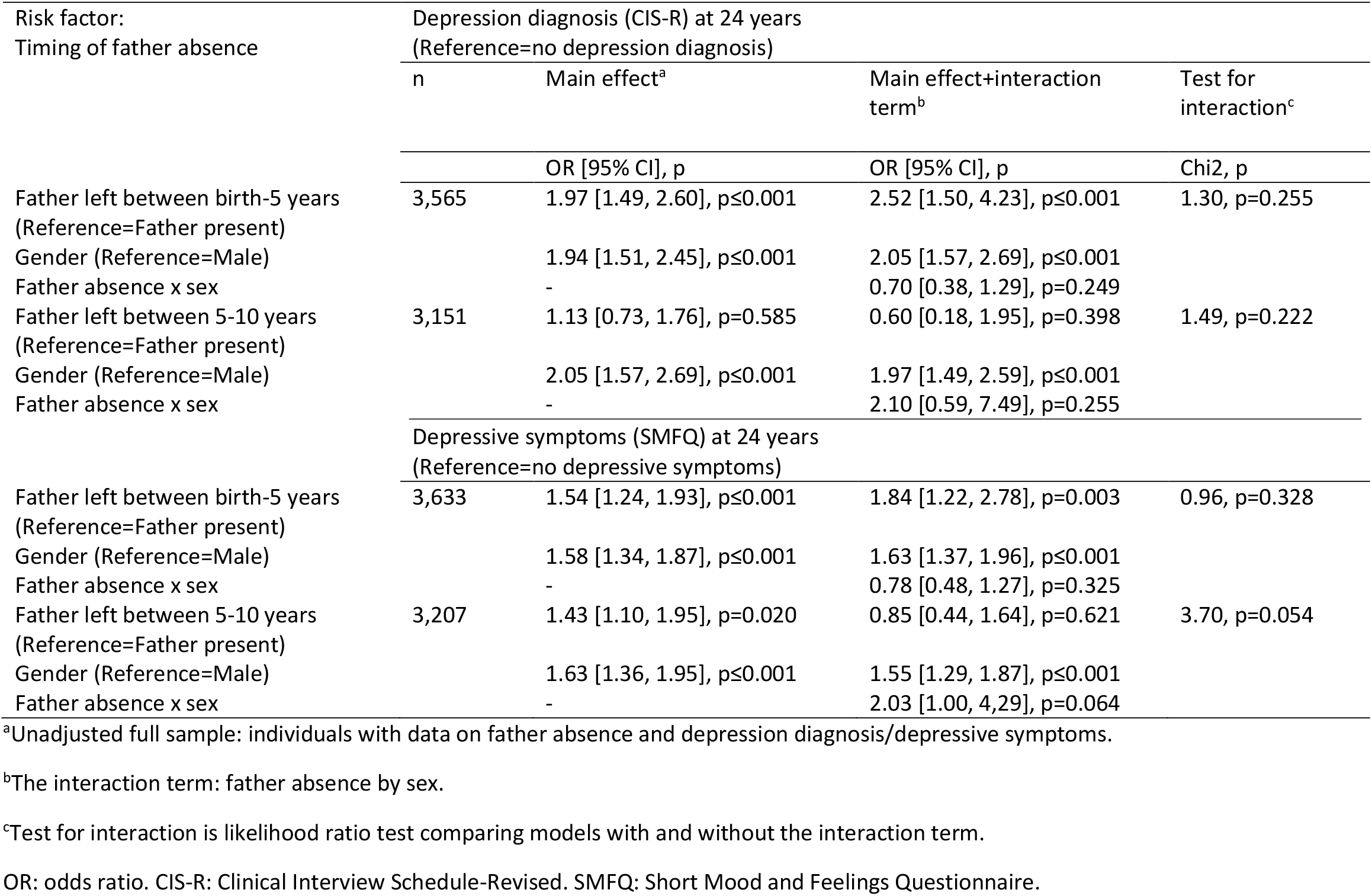
Odds ratios for [95% Cis] for the association between father absence during different periods in childhood and binary indicators of depression diagnosis and depressive symptoms at 24 years stratifying by sex in the entire sample

Second, we estimated unadjusted and adjusted models to substantiate our findings with regard to the main effects of early and middle childhood father absence on depression diagnosis and depressive symptoms at age 24 years whilst accounting for a range of confounders. Consistent with the analyses utilising the whole sample, there was strong evidence in the unadjusted model for a main effect of father absence in early childhood on depression (OR: 2.00, 95%CI 1.39, 2.89, p≤0.001; Table 2) and depressive symptoms (OR: 1.86, 95%CI 1.40, 2.46, p≤0.001; Table 2) at age 24 years. Although moderately attenuated, this association was independent of socioeconomic, maternal and familial confounders (depression: OR: 1.58, 95%CI 1.07, 2.32, p=0.021; depressive symptoms: OR: 1.52, 95%CI 1.13, 2.03, p=0.006; Table 2). In contrast, there was no evidence for a main effect of father absence in middle childhood on depression or depressive symptoms at age 24 years in the unadjusted (depression: OR: 1.13, 95%CI 0.67, 1.90, p=0.650; depressive symptoms: OR: 1.17, 95%CI 0.80, 1.72, p=0.424; Table 2) or adjusted models (depression: OR: 0.97, 95%CI 0.57, 1.65, p=0.906; depressive symptoms: OR: 1.04, 95%CI 0.70, 1.54, p=0.842; Table 2). The results from the analyses with imputed data supported our findings and led to the same overarching conclusions (Results S1, Tables S4-S5, Supplementary).

**Table 2.** Odds ratios [95% CI] for the main effect of father absence during different periods in childhood on binary indicators of depression diagnosis and depressive symptoms at 24 years

| Risk factor: | n | Depression diagnosis (CIS-R) at 24 years |  | n | Depressive symptoms (SMFQ) at 24 years |  |
| --- | --- | --- | --- | --- | --- | --- |
| Timing of father absence |  | (Reference=no depression diagnosis) |  |  | (Reference=no depressive symptoms) |  |
| Father absence<br>(Reference=Father present) |  | Unadjusted<br>OR [95% CI], p | Adjusted<sup>a</sup><br>OR [95% CI], p |  | Unadjusted<br>OR [95% CI], p | Adjusted<sup>a</sup><br>OR [95% CI], p |
| Father left between birth-5 years | 2,672 | 2.00 [1.39, 2.89],<br>p≤0.001 | 1.58 [1.07, 2.32],<br>p=0.021 | 2,694 | 1.86 [1.40, 2.46],<br>p≤0.001 | 1.52 [1.13, 2.03],<br>p=0.006 |
| Father left between 5-10 years | 2,435 | 1.13 [0.67, 1.90],<br>p=0.650 | 0.97 [0.57, 1.65],<br>p=0.906 | 2,450 | 1.17 [0.80, 1.72],<br>p=0.424 | 1.04 [1.70, 1.54],<br>p=0.842 |

*Note:* <sup>a</sup>Adjusted for antenatal indicators of socioeconomic (parental social class, financial difficulties, maternal educational attainment, homeownership), maternal (depression) and familial (parental conflict) characteristics. OR: Odds ratio. CIS-R: Clinical Interview Schedule-Revised. SMFQ: Short Mood and Feelings Questionnaire.

### Association between father absence in childhood and trajectories of depressive symptoms from childhood to early adulthood

As non-linear trajectories are difficult to interpret, the predicted mean differences between different groups at different ages are presented in Table 3, with regression coefficients from the models presented in Tables S6-S9 and Tables S10-S13, Supplementary. The predicted scores for each group at different ages are presented in Table S14, Supplementary. In the adjusted model, the overall pattern of depressive symptoms trajectories increased throughout later childhood and adolescence up to age 18 years, followed by a decrease around age 20 before rising again around age 22 years. Children exposed to father absence in early childhood, compared with presence, had similar trajectories around age 12 years, but began to have higher trajectories of depressive symptoms from around age of 14 (Figure 2). This increase in trajectories was indexed by father absence in early childhood having a greater mean difference in depressive symptoms scores from the age of 16 years (Beta^diff^ = 0.60, 95%CI 0.17, 1.02, p=0.012; Table 4), with the greatest difference observed at age 24 (Beta^diff^ = 1.06, 95%CI 0.37, 1.74, p=0.003; Table 4). Father absence in middle childhood showed a similar pattern up to the age of 16 years with evidence of a mean difference observed at this age (Beta^diff^ = 0.68, 95%CI 0.17, 1.19, p=0.008; Table 4). However, in contrast to father absence in early childhood, father absence in middle childhood was not associated with any mean differences at age 20 or 24 years respectively as the gap between father absence in middle childhood and father presence during this period trajectories narrowed into early adulthood. Results did not differ substantially with the inverse probability weighted analysis (Table S15-S16, Supplementary).

**Table 3.** Predicted mean difference in depressive symptoms scores [95% CI] at various ages for the main effect of father absence during different periods in childhood on trajectories of depressive symptoms

| Predicted Mean Differences in Depressive Symptoms Scores (SMFQ) <sup>a</sup> |  |  |  |  |
| --- | --- | --- | --- | --- |
|  | Age 12 | Age 16 | Age 20 | Age 24 |
| *Model 1* (n=6,020) |  |  |  |  |
| No Early Father Absence vs<br>Early Father Absence | 0.13 (-0.19, 0.45), p=0.426 | 0.60 (0.17, 1.02), p=0.012 | 0.57 (0.04, 1.11), p=0.048 | 1.06 (0.37, 1.75), p=0.003 |
| *Model 2* (n=5,352) |  |  |  |  |
| No Later Father Absence vs<br>Later Father Absence | 0.31 (-0.07, 0.69), p=0.190 | 0.68 (0.17, 1.19), p=0.008 | 0.37 (-0.27, 1.01), p=0.254 | 0.63 (-0.21, 1.48), p=0.143 |

*Note:* <sup>a</sup> Adjusted for antenatal indicators of socioeconomic (parental social class, financial difficulties, maternal educational attainment, homeownership), maternal (depression) and familial (parental conflict) characteristics. P-values are adjusted for false discovery rate (FDR). SMFQ: Short Mood and Feelings Questionnaire.

**Table 4.** Predicted mean difference in depressive symptoms scores [95% CI] at various ages for the main effect of father absence during different periods in childhood on trajectories of depressive symptoms stratifying by sex in the entire sample

|  | Predicted Mean Differences in Depressive Symptoms Scores (SMFQ)<sup>a</sup> |  |  |  |
| --- | --- | --- | --- | --- |
|  | Age 12 | Age 16 | Age 20 | Age 24 |
| *Model 3 (n=6020)* |  |  |  |  |
| Male No Early Father Absence vs Female No Early Father Absence | 0.40 (0.19, 0.60), p=0.0002 | 2.06 (1.80, 2.32), p=1.6x10<sup>-15</sup> | 1.56 (1.24, 1.89), p=1.6x10<sup>-15</sup> | 1.37 (0.93, 1.80), p=2.7x10<sup>-09</sup> |
| Male No Early Father Absence vs Male Yes Early Father Absence | 0.01 (-0.47, 0.45), p=0.974 | 0.52 (-0.11, 1.15), p=0.14 | 0.25 (-0.61, 1.11), p=0.624 | 2.11 (0.95, 3.27), p=0.0009 |
| Male No Early Father Absence vs Female Yes Early Father Absence | 0.65 (0.21, 1.10), p=0.007 | 2.68 (2.13, 3.24), p=1.6x10<sup>-15</sup> | 2.25 (1.56, 2.94), p=1.1x10<sup>-09</sup> | 1.87 (0.99, 2.74), p=0.00009 |
| Female No Early Father Absence vs Male Yes Early Father Absence | 0.40 (-0.06, 0.87), p=0.12 | 1.54 (0.91, 2.16), p=4.8x10<sup>-06</sup> | 1.32 (0.47, 2.17), p=0.004 | 0.74 (0.40, 1.88), p=0.253 |
| Female No Early Father Absence vs Female Yes Early Father Absence | 0.26 (-0.19, 0.70), p=0.294 | 0.63 (0.07, 1.18), p=0.044 | 0.69 (0.01, 1.36), p=0.072 | 0.50 (-0.35, 1.36) p=0.294 |
| Male Yes Early Father Absence vs Female Yes Early Father Absence | 0.66 (0.06, 1.26), p=0.049 | 2.16 (1.37, 2.95) p=3.3x10<sup>-07</sup> | 2.00 (0.96, 3.05), p=0.0002 | 0.24 (-1.13, 1.61), p=0.762 |
| *Model 4 (n=5352)* |  |  |  |  |
| Male No Later Father Absence vs Female No Later Father Absence | 0.38 (0.18, 0.59), p=0.0006 | 1.94 (1.67, 2.20), p=1.6x10<sup>-15</sup> | 1.45 (1.12, 1.79), p=1.6x10<sup>-15</sup> | 1.28 (0.84, 1.72), p=3.6x10<sup>-08</sup> |
| Male No Later Father Absence vs Male Yes Later Father Absence | 0.28 (-0.27, 0.94), p=0.362 | 0.34 (-0.44, 1.11), p=0.429 | 0.67 (-0.38, 1.72), p=0.253 | 0.06 (-1.43, 1.54), p=0.939 |
| Male No Later Father Absence vs Female Yes Later Father Absence | 0.80 (0.27, 1.33), p=0.006 | 3.20 (2.55, 3.85), p=1.6x10<sup>-15</sup> | 2.37 (1.56, 3.18), p=3.7x10<sup>-08</sup> | 2.34 (1.26, 3.41), p=0.00005 |
| Female No Later Father Absence vs Male Yes Later Father Absence | 0.10 (-0.46, 0.65), p=0.759 | 2.28 (1.50, 3.05), p=3.5x10<sup>-08</sup> | 2.12 (1.08, 3.16), p=0.0001 | 1.34 (-0.14, 2.81), p=0.105 |
| Female No Later Father Absence vs Female Yes Later Father Absence | 0.42 (-0.11, 0.95), p=0.162 | 1.26 (0.61, 1.91), p=0.0002 | 0.92 (0.12, 1.72), p=0.038 | 1.06 (0.00, 2.12), p=0.076 |
| Male Yes Later Father Absence vs Female Yes Later Father Absence | 0.52 (-0.22, 1.25), p=0.215 | 3.53 (2.56, 4.51), p=6.2x10<sup>-12</sup> | 3.04 (1.77, 4.32), p=8.7x10<sup>-06</sup> | 2.39 (0.62, 4.17), p=0.013 |

*Note:* <sup>a</sup>Adjusted for antenatal indicators of socioeconomic (parental social class, financial difficulties, maternal educational attainment, homeownership), maternal (depression) and familial (parental conflict) characteristics. P-values are adjusted for false discovery rate (FDR). SMFQ: Short Mood and Feelings Questionnaire.

**Figure 2.**
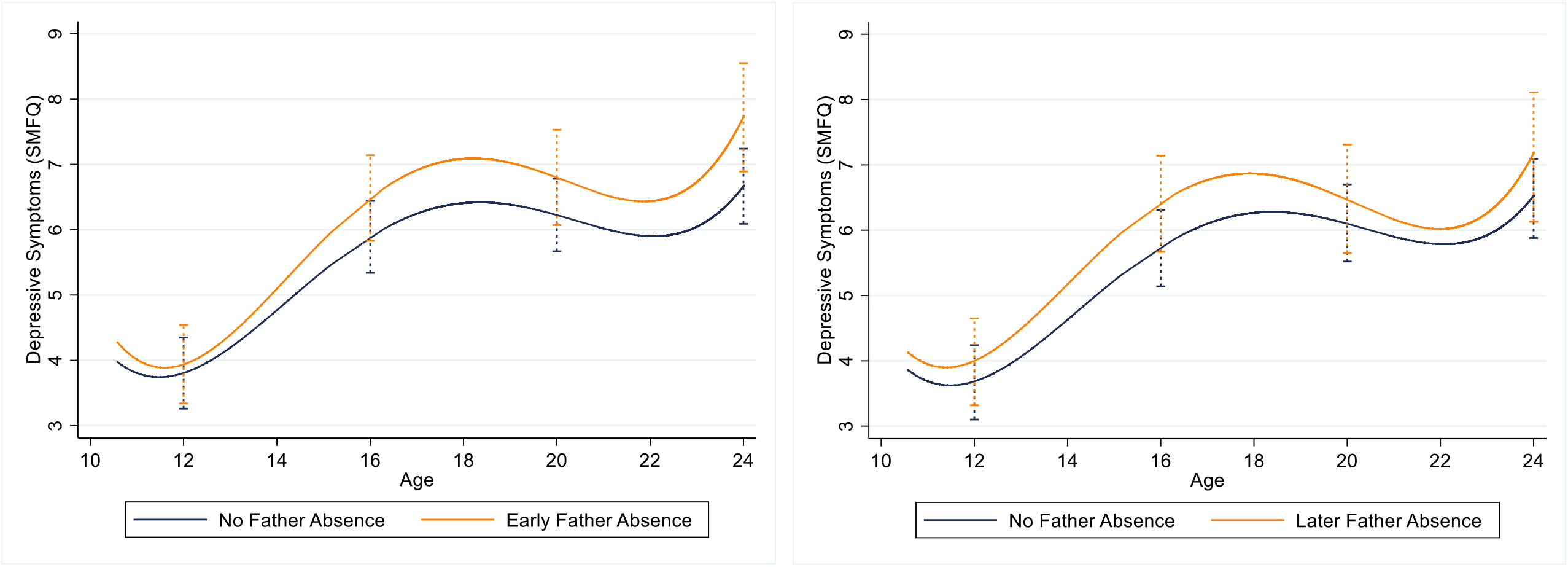
Main effect of father absence early in childhood (birth-5 years; left) and father absence in middle childhood (5-10 years; right) on predicted trajectories of depressive symptoms across childhood, adolescence and young adulthood

There was clear evidence that females had higher trajectories of depressive symptoms compared to males across adolescence and young adulthood (Figure 3). Females exposed to father absence in early and middle childhood had the highest trajectories, with males whose fathers were present across childhood displaying the lowest trajectories. Males whose fathers were absent in early childhood had similar trajectories to those whose fathers were present until the age 24 years, followed by a steep rise in depressive symptoms with a greater mean difference in depressive symptoms scores at age 24 years (Beta^diff^ = 2.11, 95%CI 0.95, 3.27, p=0.009; Table 4). Compared with females whose fathers were present, those whose fathers were absent in early childhood had higher depressive symptoms trajectories throughout adolescence, but this difference narrowed by age 24 (Beta^diff^ = 0.50, 95%CI -0.35, 1.36, p=0.294; Table 4). Compared with males, females who were exposed to father absence in early childhood had higher depressive symptom trajectories across late childhood and adolescence, but the sharp rise in depressive symptoms in adulthood for males exposed to early father absence resulted in trajectories that were non-discriminant at age 24 years (Beta^diff^ = 0.24, 95%CI -1.13, 1.61, p=0.762; Table 4).

**Figure 3.**
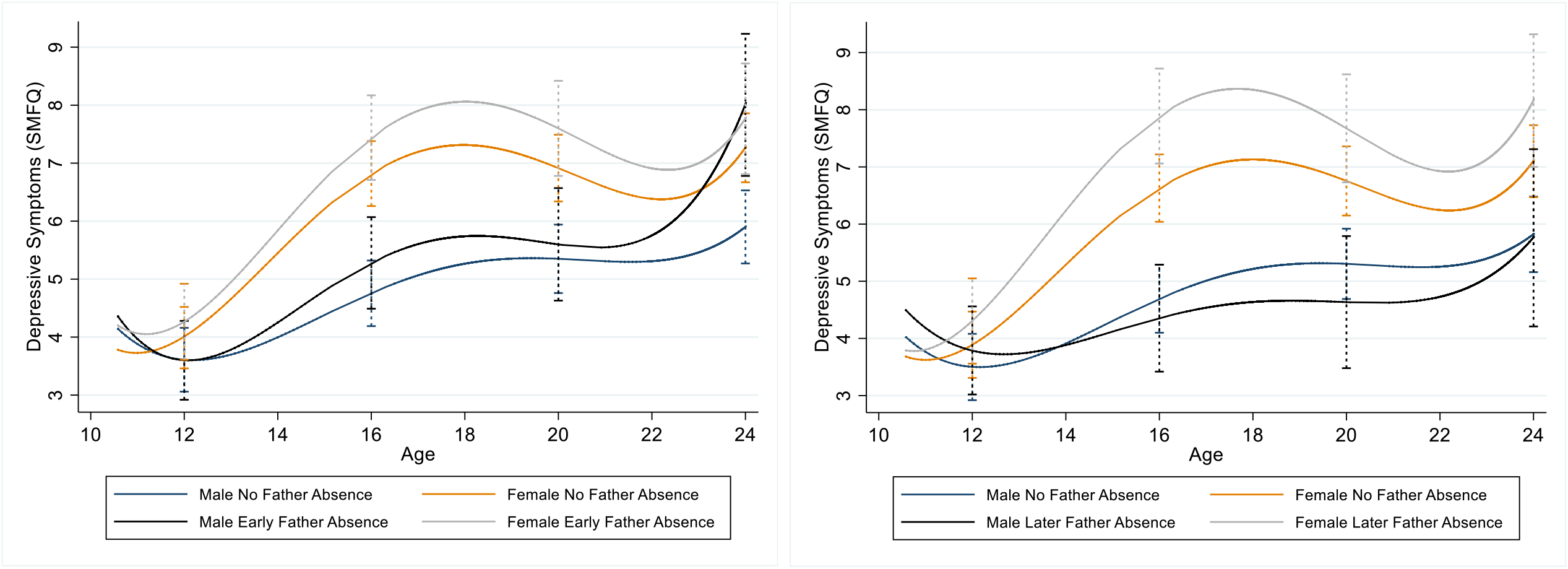
Main effect of early father absence (left; birth-5 years) and later father absence in middle childhood (right; 5-10 years) on predicted trajectories of depressive symptoms across childhood, adolescence and young adulthood, stratified by gender

There was no steep rise in depression trajectories in adulthood observed for males whose father was absent in middle childhood, with little difference in depressive symptoms scores at age 24 years between father present males (Beta^diff^ = 0.06, 95%CI -1.43, 1.54, p=0.939; Table 4). In contrast, males with later father absence had the lowest trajectories of all, although these were non-discriminant from males whose fathers were present. There was strong evidence that females exposed to father absence in middle childhood had higher depression trajectories than their unexposed female counterparts at age 16 (Beta^diff^ = 1.26, 95%CI 0.61, 1.91, p=0.002) and 20 years (Beta^diff^ = 0.92, 95%CI 0.12, 1.72, p=0.038), though this difference had narrowed somewhat by age 24 years (Beta^diff^ = 1.06, 95%CI 0.00, 2.12, p=0.076; Table 4). Similarly to early father absence model, females whose fathers were absent in middle childhood had higher depressive symptoms trajectories compared to males whose fathers left in middle childhood across all ages except age 12 years (Beta^diff^ = 0.52, 95%CI -0.22, 1.25, p=0.215; Table 4).

## Discussion

### Main findings

In this large population-based study we found that early but not middle childhood father absence was associated with increased odds of offspring depression at age 24. We found no evidence for a sex difference in this relationship. Father absence in both early and middle childhood was associated with increased odds of depressive symptoms at age 24 years, with some evidence of this relationship being stronger for females than males. Consistently, father absence in early childhood was strongly associated with increased odds of offspring depression and depressive symptoms at age 24 years in the unadjusted and fully adjusted models. Although moderately attenuated, these associations were independent of socioeconomic, maternal and familial confounding factors. In contrast, there was no evidence to suggest that father absence in middle childhood was associated with increased odds of either offspring depression or depressive symptoms at age 24 years in the unadjusted and fully adjusted models.

Modelling developmental trajectories of depressive symptoms enabled us to track the effects of biological father absence during different periods in childhood on offspring mental health over time, providing insights into whether these effects persist or diminish with age. The overall pattern of depressive symptoms was consistent with that observed in previous studies, with initially low levels of depressive symptoms in late childhood increasing through adolescence up to age 18, followed by a decline before rising again at the age of 22.^34^ There was clear evidence that females, on average, had much worse trajectories compared to father absent and father present males across adolescence and young adulthood, with males whose fathers were present throughout childhood displaying the lowest trajectories across development. These findings are consistent with previous research suggesting that females are more likely than males to have increasing trajectories of depressive symptoms from childhood through late adolescence^35,36^ and early adulthood.^37^

Father absence in early childhood was associated with higher depressive symptom trajectories across adolescence with the greatest difference in mean depressive symptoms scores between father absent and present groups observed at age 24 years. This suggests that there is a persistent association between early father absence and depressive symptoms in late adolescence and young adulthood. Offspring whose fathers were absent during middle childhood had similar trajectories until mid-adolescence. However, in contrast to the effects of early father absence, the gap between offspring whose father were absent and present in middle childhood tended to narrow into early adulthood, suggesting that there may be differential mechanisms underlying timing effects of early father absence. In line with the family stress model,^38^ early childhood adverse experiences, including parental separation and divorce, may trigger acute and chronic economic hardship,^39^ parental conflict and psychological distress,^39^ and disrupted parenting that put children at risk of long-term psychological problems.^40^ Patterns of non-resident father contact may also be an important mechanism underlying differential effects of biological father absence on depressive symptoms. Evidence suggests that early life family dissolution is associated with a gradual decline in the frequency of contact and involvement between the non-resident father and the child over time.^41^ Arguably, fathers who leave when the child is older had more opportunities to become involved in child rearing during the early years, and involved fathers are more likely to remain in contact with their children,^42,43^ reducing the risk of adverse mental health in offspring of separated parents.^44^

There was also evidence of sex specific effects of biological father absence on trajectories of offspring depressive symptoms. Specifically, females whose fathers were absent in early childhood had higher depressive symptoms trajectories throughout adolescence compared to their father present counterparts, but this difference had narrowed by age 24. These findings are in line with previous research emphasising adverse effects of parental divorce^41^ and biological father absence on offspring depressive symptoms, particularly for females.^7^ Similar patterns of depressive symptoms trajectories in females from divorced compared to nondivorced families have been previously reported.^16,45,46^ The rise in female depressive symptoms during early to middle adolescence may coincide with early pubertal transition, particularly in the context of biological father absence,^47^ and a range of other developmental and psychosocial stressors and transitions that may explain increased risk of emotional distress.^48,49^ There is strong evidence for an association between father absence and early timing of menarche in girls,^50^ but it is unclear whether father absence affects pubertal timing in boys.^51^ The decline in trajectories during transition to young adulthood in females who experienced early childhood biological father absence is noteworthy and requires further research to understand underlying mechanisms. It has been previously suggested that increased levels of autonomy and decreased reliance on parental support,^52^ relational interdependency and formation of intimate pair bonds^53^ characteristic of this developmental period, as well as availability of wider support networks,^54^ may decrease risk for psychological distress in females.

A different pattern of depressive symptoms trajectories was observed for males whose fathers were absent in early childhood. Specifically, these males had similar trajectories to their father present counterparts until early adulthood where early father absent males experienced a steep rise in depressive symptoms between the ages of 22-24. Vulnerabilities such as renewed sense of parental loss and lack of appropriate male role models or models of long-term intimate adult relationships^41^ at the time when the importance of interpersonal relationships becomes more salient may be the underlying exploratory mechanisms.

Females whose fathers were absent in middle childhood had higher depressive symptoms trajectories across middle adolescence and into young adulthood compared to their father present counterpart, with the widest gap around ages 16-20 years, which could be a key developmental period to intervene. This finding is consistent with our full sample analyses suggesting that negative effects of father absence in middle childhood are stronger for females compared to males. In contrast, males whose fathers were absent in middle childhood had the lowest trajectories of all, although these were non-discriminant from their father present counterparts. Based on existing literature, it is possible that that rising trajectories for girls may be in line with the overall pattern of higher lifetime trajectories for females than for males. Still, it is unclear why only males and not females show declining trajectories in response to middle childhood father absence. It may be that older children have more mature cognitive and emotional resources to cope with the changing nature of father-child relationship,^55^ while sex differences in experiencing and coping with interpersonal emotional distress^56^ could explain the more adverse effect of biological father absence for females than males. There is also some evidence that fathers spend more time with their sons than daughters during marriage,^3^ a pattern that may persist regarding initiation, frequency and quality of contact with the non-resident fathers following family dissolution.^57,58^ Although inconsistent, evidence suggests that not only sons are more likely to be in contact with their non-resident fathers more than daughters, they also perceive the quality of the relationship to be better.^59,60^

### Strengths and limitations

The main strengths are the population-based design with prospectively collected data and repeated measures of depressive symptoms and biological father absence, which reduces the possibility of selection and recall bias. The availability of these data enabled us to model differential effects of biological father absence at distinct periods in childhood on clinical diagnosis of depression and depressive symptoms in early adulthood, as well as longitudinal trajectories of mental health from childhood to early adulthood. Furthermore, the advantage of multiple assessments of depressive symptoms over time (from late childhood to adolescence, across adolescence, adolescence to young adulthood) enabled us to examine at which ages father absence may exert its most prominent effect on offspring trajectories of depressive symptoms and whether the effects were short lived or persistent across development. The availability of rich covariable information enabled us to control for a number of social and contextual confounding factors preceding father departure, including parental conflict, which is an important risk factor for both family dissolution^60^ and adverse offspring mental health.^61^ Lack of adjustment for preceding parental conflict has been argued as a limitation of existing research on the effect of biological father absence.^5^ Although we adjusted for a range of confounding factors to minimise the possibility of confounding bias, we cannot fully eliminate the possibility of unmeasured and residual confounding. We were also unable to account for the frequency of contact and quality of the relationship between the non-resident father and the child, as well as presence of an alternative father figure. The limitation of our study is sample attrition, which is similar of that observed in other population-based studies.^19,20^ Attrition may undermine validity of our findings, given that participants from lower socio-economic background with higher prevalence of father absence and depression were somewhat under-represented in our study. We addressed bias associated with selection attrition by controlling for a range of factors known to predict missingness and by imputing missing data in outcome and confounders. The pattern of missing data and imputed analyses suggested that attrition may have led to the underestimation of main effects in complete case analyses. Another potential limitation is longitudinal measurement invariance as explored in previous research,^31^ however, given that depression trajectories are based on a summary score and we are not focusing on specific components or constructs of depression, this impact is likely to be limited.

### Conclusions and implications of the research

Our findings suggest that the adverse effects of early childhood biological father absence on offspring mental health may persist across adolescence and into early adulthood. Examining sex and timing effects of father absence on offspring depression at distinct developmental periods, as well as longitudinal trajectories across the life course provides a more nuanced understanding of how these effects change with age and their potential differential impact by sex. Such insights are important for targeted interventions. Further research is needed to examine whether these associations are causal to strengthen present findings, as well as efforts to provide insights into mechanisms underlying sex and timing effects of father absence on depressive symptoms to facilitate development of targeted interventions.

## Data Availability

ALSPAC data are available through a system of managed open access. The study website contains details of all the data that is available through a fully searchable data dictionary and variable search tool data dictionary. The application steps for ALSPAC data access are highlighted below.
1. Please read the ALSPAC access policy, which describes the process of accessing the data in detail, and outlines the costs associated with doing so.
2. You may also find it useful to browse the fully searchable research proposals database, which lists all research projects that have been approved since April 2011.
3. Please submit your research proposal for consideration by the ALSPAC Executive Committee. You will receive a response within 10 working days to advise you whether your proposal has been approved.
If you have any questions about accessing data, please.

## Funding statement

The UK Medical Research Council and Wellcome (Grant ref: 217065/Z/19/Z) and the University of Bristol provide core support for ALSPAC. This publication is the work of the authors and will serve as guarantors for the contents of this paper. A comprehensive list of grants funding is available on the ALSPAC website (http://www.bristol.ac.uk/alspac/external/documents/grant-acknowledgements.pdf).

This research was funded in whole by the Wellcome Trust Research Fellowship in Humanities and Social Science (Grant ref: 212664/Z/18/Z) awarded to Dr Culpin. For the purpose of Open Access, the author has applied a CC BY public copyright licence to any Author Accepted Manuscript version arising from this submission. Dr Pearson was supported by the European Research Commission Grant (Grant ref: 758813 MHINT). Dr Kwong is supported by an Economic Social Research Council Postdoctoral Fellowship (Grant ref: ES/V011650/1). This study was also supported by the NIHR Biomedical Research Centre at the University Hospitals Bristol and Weston NHS Foundation Trust and the University of Bristol. This publication is the work of the authors who will serve as guarantors for the contents of this paper. The views expressed in this publication are those of the author(s) and not necessarily those of the NHS, the National Institute for Health Research.

## Conflicts of Interest

None.

## Ethical standards

Informed consent for the use of data collected via questionnaires and clinics was obtained from participants following the recommendations of the ALSPAC Ethics and Law Committee at the time. The authors assert that all procedures contributing to this work comply with the ethical standards of the relevant national and institutional committees on human experimentation and with the Helsinki Declaration of 1975, as revised in 2008.

## Acknowledgements

We are extremely grateful to all the families who took part in this study, the midwives for their help in recruiting them, and the whole ALSPAC team, which includes interviewers, computer and laboratory technicians, clerical workers, research scientists, volunteers, managers, receptionists and nurses.

## Notes

### Competing Interest Statement

The authors have declared no competing interest.

### Author Declarations

Ethical approval for the study was obtained from the ALSPAC Ethics and Law Committee and the Local Research Ethics Committees.

## References

1. ONS (2013). Retrieved from Divorces in England and Wales - Office for National Statistics (ons.gov.uk).

2. Crawford, C., Goodman, A., Greaves, E. & Joyce, R. (2011). Cohabitation, marriage, relationship stability and child outcomes: an update. Retrieved from http://www.ifs.org.uk/comms/comm120.pd

3. Kalmijn, M. (2015). Father-child relations after divorce in four European countries: Patterns and determinants. Comparative Population Studies, 40(3). doi: https://doi.org/10.12765/CPoS-2015-10.

4. Amato, P. R. (2001). Children of divorce in the 1990s: an update of the Amato and Keith (1991) meta-analysis. Journal of Family Psychology, 15(3), 355–370.

5. Amato, P. R., & Keith, B. (1991). Parental divorce and the well-being of children: a meta-analysis. Psychological Bulletin, 110(1), 26–46.

6. McLanahan, S., Tach, L., & Schneider, D. (2013). The causal effects of father absence. Annual Review of Sociology, 39, 399–427.

7. Culpin, I., Heron, J., Araya, R., Melotti, R., & Joinson, C. (2013). Father absence and depressive symptoms in adolescence: findings from a UK cohort. Psychological Medicine, 43(12), 2615–2626.

8. Oldehinkel, A. J., Ormel, J., Veenstra, R., De Winter, A. F., & Verhulst, F. C. (2008). Parental divorce and offspring depressive symptoms: Dutch developmental trends during early adolescence. Journal of Marriage and Family, 70(2), 284–293.

9. Sigle-Rushton, W., & McLanahan, S. (2004). Father absence and child wellbeing: a critical review (pp. 116–55). kNew York: Russell Sage Foundation.

10. Ermisch, J. F., & Francesconi, M. (2001). Family structure and children’s achievements. Journal of population economics, 14(2), 249–270.

11. Rudolph, K. D. (2002). Gender differences in emotional responses to interpersonal stress during adolescence. Journal of adolescent health, 30(4), 3–13.

12. Størksen, I., Røysamb, E., Moum, T., & Tambs, K. (2005). Adolescents with a childhood experience of parental divorce: a longitudinal study of mental health and adjustment. Journal of adolescence, 28(6), 725–739.

13. Morrison, D. R., & Cherlin, A. J. (1995). The divorce process and young children’s well-being: a prospective analysis. Journal of Marriage and the Family, 800–812.

14. Sameroff, A. J. (2000). Dialectical processes in developmental psychopathology. In Handbook of Developmental Psychopathology (pp. 23–40). Springer, Boston, MA.

15. Herle, M., Micali, N., Abdulkadir, M., Loos, R., Bryant-Waugh, R., Hübel, C., Bulik, C. M., & De Stavola, B. L. (2020). Identifying typical trajectories in longitudinal data: modelling strategies and interpretations. European Journal of Epidemiology, 35(3), 205–222.

16. Ge, X., Natsuaki, M. N., & Conger, R. D. (2006). Trajectories of depressive symptoms and stressful life events among male and female adolescents in divorced and nondivorced families. Developmental Psychopathology, 18(1), 253–273. doi:10.1017/S0954579406060147

17. Cherlin, A. J., Chase-Lansdale, P. L., & McRae, C. (1998). Effects of parental divorce on mental health throughout the life course. American Sociological Review, 239–249.

18. Costello, D. M., Swendsen, J., Rose, J. S., & Dierker, L. C. (2008). Risk and protective factors associated with trajectories of depressed mood from adolescence to early adulthood. Journal of Consulting and Clinical Psychology, 76(2), 173–183. doi:10.1037/0022-006X.76.2.173.

19. Boyd, A., Golding, J., Macleod, J., Lawlor, D. A., Fraser, A., Henderson, J., Molloy Ness, A., Ring, S., & Davey Smith, G. (2013). Cohort Profile: The ‘Children of the 90s’: the index offspring of the Avon Longitudinal Study of Parents and Children. International Journal of Epidemiology, 42, 111–127.

20. Fraser, A., Macdonald-Wallis, C., Tilling, K., Boyd, A., Golding, J., Davey Smith, G., Henderson, J., Macleod, J., Molloy, L., Ness, A., Ring, S., Nelson, S. M., & Lawlor, D. A. (2012). Cohort profile: the Avon Longitudinal Study of Parents and Children: ALSPAC mothers cohort. International Journal of Epidemiology, 42, 97–110.

21. Northstone, K., Lewcock, M., Groom, A., Boyd, A., Macleod, J., Timpson, N.J., Wells, N. (2019). The Avon Longitudinal Study of Parents and Children (ALSPAC): an updated on the enrolled sample of index children in 2019. Wellcome Open research; 4:51 (https://doi.org/10.12688/wellcomeopenres.15132.1)

22. Harris, P.A., Taylor, R., Thielke, R., Payne, J.N, Gonzalez, JG. (2009). Conde, Research Electronic Data capture (REDCap) – A metadata-driven methodology and workflow process for providing translational research informatics support. Journal of Biomedical Informatics, 42, 377–381.

23. Lewis, G., Pelosi, A. J., Araya, R., & Dunn, G. (1992). Measuring psychiatric disorder in the community: a standardized assessment for use by lay interviewers. Psychological Medicine, 22, 465–486.

24. Thapar, A., & McGuffin, P. (1998). Validity of the shortened Mood and Feelings Questionnaire in a community sample of children and adolescents: a preliminary research note. Psychiatry Research, 81(2), 259–268.

25. Angold, A., Costello, E.J., Messer, S.C., Pickles, A., Winder, F., & Silver, D. (1995). Development of a short questionnaire for use in epidemiological studies of depression in children and adolescents. International Journal of Methods in Psychiatric Research, 5, 237–249.

26. Turner, N., Joinson, C., Peters, T. J., Wiles, N., & Lewis, G. (2014). Validity of the Short Mood and Feelings Questionnaire in late adolescence. Psychological Assessment, 26(3), 752.

27. Patton, G. C, Coffey, C., Posterino, M., Carlin, J. B., Wolfe, R., & Bowes, G. (1999). A computerised screening instrument for adolescent depression: population-based validation and application to a two-phase case-control study. Social Psychiatry and Psychiatric Epidemiology, 34, 166–172.

28. Shaffer, D., Fisher, P., Lucas, C. P., Dulcan, M. K., & Schwab-Stone, M. E. (2000). NIMH Diagnostic Interview Schedule for Children Version IV (NIMH DISC-IV): description, differences from previous versions, and reliability of some common diagnoses. Journal of the American Academy of Child & Adolescent Psychiatry, 39(1), 28–38.

29. Kwong, A. S., Manley, D., Timpson, N. J., Pearson, R. M., Heron, J., Sallis, H., Stergiakouli, E., Davis, O. S. P., & Leckie, G. (2019). Identifying critical points of trajectories of depressive symptoms from childhood to young adulthood. Journal of Youth and Adolescence, 48(4), 815–827. https://link.springer.com/article/10.1007/s10964-018-0976-5

30. Cox, J. L., Holden, J. M., & Sagovsky, R. (1987). Detection of postnatal depression: development of the 10-item Edinburgh Postnatal Depression Scale. British Journal of Psychiatry, 150, 782–786.

31. Kwong, A. S., Morris, T. T., Pearson, R. M., Timpson, N. J., Rice, F., Stergiakouli, E., & Tilling, K. (2021). Polygenic risk for depression, anxiety and neuroticism are associated with the severity and rate of change in depressive symptoms across adolescence. Journal of Child Psychology and Psychiatry. Doi: https://doi.org/10.1111/jcpp.13422.

32. Stata v.15/MP. StataCorp., USA; New in Stata 15 | Stata.

33. Leckie, G., & Charlton, C. (2013). runmlwin - A Program to Run the MLwiN Multilevel Modelling Software from within Stata. Journal of Statistical Software, 52 (11),1-40. (do).

34. Kwong, A. S. (2019). Examining the longitudinal nature of depressive symptoms in the Avon Longitudinal Study of Parents and Children (ALSPAC). Wellcome open research, 4.

35. Dekker, M. C., Ferdinand, R. F., Van Lang, N. D., Bongers, I. L., Van Der Ende, J., & Verhulst, F. C. (2007). Developmental trajectories of depressive symptoms from early childhood to late adolescence: gender differences and adult outcome. Journal of Child Psychology and Psychiatry, 48(7), 657–666.

36. Lewis, A. J., Sae-Koew, J.H. Toumbourou, J.W., & Rowland, B. (2020). Gender differences in trajectories of depressive symptoms across childhood and adolescence: a multi-group growth mixture model. Journal of Affective Disorders, 260, 463–472.

37. Adkins, D. E., Wang, V., Dupre, M. E., Van den Oord, E. J., & Elder, G. H. (2009). Structure and stress: trajectories of depressive symptoms across adolescence and young adulthood. Social Forces, 88, 31–60.

38. Conger, R. D., Elder Jr, G. H., Lorenz, F. O., Conger, K. J., Simons, R. L., Whitbeck, L. B., Huck, S., & Melby, J. N. (1990). Linking economic hardship to marital quality and instability. Journal of Marriage and the Family, 52(3), 643–656.

39. Conger, R. D., Conger, K. J., & Martin, M. J. (2010). Socioeconomic status, family processes, and individual development. Journal of Marriage and Family, 72(3), 685–704.

40. Masarik, A. S., & Conger, R. D. (2017). Stress and child development: a review of the Family Stress Model. Current Opinion in Psychology, 13, 85–90.

41. Chase-Lansdale, P. L., Cherlin, A. J., & Kiernan, K. E. (1995). The long-term effects of parental divorce on the mental health of young adults: A developmental perspective. Child Development, 66(6), 1614–1634.

42. De Graaf, P. M., & Fokkema, T. (2007). Contacts between divorced and non-divorced parents and their adult children in the Netherlands: An investment perspective. European Sociological Review, 23(2), 263–277.

43. Westphal, S. K., Poortman, A. R., & Van Der Lippe, T. (2014). Non-resident father–child contact across divorce cohorts: The role of father involvement during marriage. European Sociological Review, 30(4), 444–456.

44. Flouri, E. (2006). Non-resident fathers’ relationships with their secondary school age children: Determinants and children’s mental health outcomes. Journal of Adolescence, 29(4), 525–538.

45. Hankin, B. L. (2009). Development of sex differences in depressive and co-occurring anxious symptoms during adolescence: Descriptive trajectories and potential explanations in a multiwave prospective study. Journal of Clinical Child & Adolescent Psychology, 38(4), 460–472.

46. Stapinski, L. A., Montgomery, A. A., Heron, J., Jerrim, J., Vignoles, A., & Araya, R. (2013). Depression symptom trajectories and associated risk factors among adolescents in Chile. Plos one, 8(10), e78323.

47. Culpin, I., Heron, J., Araya, R., Melotti, R., Lewis, G., & Joinson, C. (2014). Father absence and timing of menarche in adolescent girls from a UK cohort: the mediating role of maternal depression and major financial problems. Journal of Adolescence, 37(3), 291–301.

48. Ellis, B. J. (2004). Timing of pubertal maturation in girls: an integrated life history approach. Psychological Bulletin, 130(6), 920.

49. Mendle, J., Turkheimer, E., & Emery, R. E. (2007). Detrimental psychological outcomes associated with early pubertal timing in adolescent girls. Developmental Review, 27(2), 151–171.

50. Culpin, I., Heron, J., Araya, R., Melotti, R., Lewis, G., & Joinson, C. (2014). Father absence and timing of menarche in adolescent girls from a UK cohort: the mediating role of maternal depression and major financial problems. Journal of Adolescence, 37(3), 291–301.

51. Belsky, J., Vandell, D. L., Burchinal, M., Clarke-Stewart, K. A., McCartney, K., Owen, M. T., & NICHD Early Child Care Research Network. (2007). Are there long-term effects of early child care? Child Development, 78(2), 681–701.

52. Lapsley, D., & Woodbury, R.D. (2016). Social cognitive development in emerging adulthood. In The Oxford Handbook of Emerging Adulthood. Arnett, J.J., Ed.; Oxford University Press: New York, NY, USA.

53. Gutman, L. M., & Sameroff, A. J. (2004). Continuities in depression from adolescence to young adulthood: Contrasting ecological influences. Development and Psychopathology, 16(4), 967–984.

54. Gorrese, A., & Ruggieri, R. (2012). Peer attachment: A meta-analytic review of gender and age differences and associations with parent attachment. Journal of Youth and Adolescence, 41(5), 650–672.

55. Kelly, J. B. (2003). Changing perspectives on children’s adjustment following divorce: A view from the United States. Childhood, 10(2), 237–254.

56. Seiffge-Krenke, I., & Stemmler, M. (2002). Factors contributing to gender differences in depressive symptoms: A test of three developmental models. Journal of youth and adolescence, 31(6), 405–417.

57. Manning, W. D., & Smock, P. J. (1999). New families and nonresident father-child visitation. Social Forces, 78(1), 87–116.

58. Cheadle, J. E., Amato, P. R., & King, V. (2010). Patterns of non-resident father contact. Demography, 47(1), 205–225.

59. De Wit, E., Louw, D., & Louw, A. (2014). Patterns of contact and involvement between adolescents and their non-resident fathers. Social Work, 50(1), 116–133.

60. Kelly, J. B. (2000). Children’s adjustment in conflicted marriage and divorce: A decade review of research. Journal of the American Academy of Child & Adolescent Psychiatry, 39(8), 963–973.

61. Repetti, R. L., Taylor, S. E., & Seeman, T. E. (2002). Risky families: family social environments and the mental and physical health of offspring. Psychological Bulletin, 128(2), 330.

