## Supplementary Material for "Father absence and trajectories of offspring mental health across adolescence and young adulthood: findings from a UK-birth cohort"

**Methods S1**

*Measures*

*Exposure: biological father absence in childhood*

Absence of the biological father was assessed through maternal self-reported questionnaires at regular intervals since the birth of the study child (1 year 7 months, 2 years 7 months, 3 years 9 months, 7 years, 8 years, and 10 years). To examine possible differential effect of father absence timing on offspring depression, data on biological father absence were divided into two distinct age periods to capture father absence during early (birth to 5 years) and middle (5 to 10 years) childhood.

*Outcome: offspring depression and depressive symptoms*

Offspring depressive symptoms were assessed using the Short Mood and Feelings Questionnaire (SMFQ),^1^ a 13-item scale relating to the occurrence of low mood during past two weeks, on nine occasions between ages 10 and 24 years via postal questionnaires or in research clinics. Scores for the individual items (values between 0-2) were summed up to produce a summary score ranging from 0-26. A binary variable was derived with a cut-off point of ≥10, shown to have high sensitivity and specificity for diagnosed depression,^2^ to identify individuals with depressed mood at age 24 years (*versus* no depressed mood). Details on the use of SMFQ in longitudinal models, including correlations between ages and inter-item reliability, have been reported previously.^3^

*Statistical analysis: trajectories of depressive symptoms*

We examined trajectories of depressive symptoms (as continuous SMFQ scores) between the ages of 10 and 24 years among those whose fathers were absent/present and then again separately for males and females using multilevel growth-curve models with random intercepts and random slopes to examine how depressive symptoms changed across development for each father absence model. We ran both adjusted and unadjusted models to match the depression diagnosis analyses. We interacted the age terms with the main effect of early (birth-5 years) and later (5-10 years) father absence across two separate models, followed by age terms interacted with gender across two separate models to create trajectories for each risk group. Higher order polynomials have previously been used in multilevel growth curve modelling on this data supporting this approach.^4^ Due to the complexity of interpreting higher order polynomial estimates from growth curves, we calculated the predicted mean scores for each trajectory at ages 12, 16, 20 and 24 years, followed by calculating the predicted mean difference at the varying ages between trajectories (e.g., father present at age 12 years versus early father absence at age 12 years; or male father present at age 16 years versus female later father absence at age 16 years). These time periods were chosen to delineate different developmental stages, such as early and middle adolescence and early adulthood. All analyses were conducted using Stata v.15/MP (StataCorp., USA),^5^ including trajectories estimated using the user-written *runmlwin* command (that calls the standalone multilevel modelling package MLwiN v3.04 ([www.cmm.bristol.ac.uk/MLwiN/index.shtml](http://www.cmm.bristol.ac.uk/MLwiN/index.shtml)).^6^

*Missing data: multiple imputation, full information likelihood and inverse probability weighted analysis*

We compared characteristics of missing children with those who comprised the study sample (Table S3). We accounted for the loss to follow-up by repeating our analyses on the imputed dataset. We imputed missing data because ignoring those with missing data can result in bias by assuming that data are Missing Completely at Random (MCAR).^7^ We used Multivariable Imputation by Chained Equations (MICE)^8^ to impute missing data in outcome and confounders (father absence birth-5 years: n=10,946; father absence 5-10 years: n=9,003) using the *ice* command in Stata v.15.1/MP (Stata.Corp., Texas, USA).^5^ ALSPAC provides a wealth of rich, prospectively collected data on a range of sociodemographic and mental health variables (multiple measures between ages 10 and 24 years), which enabled us to account for missing data in outcome and confounders, as well as factors that explain missingness, which is valid under the Missing-At-Random assumption (MAR).^7^ The imputation model was fully compatible with the main analyses. Using binary, ordinal logistic and regression models as appropriate, 100 imputed datasets by 10 cycles of regression switching were generated. Monte-Carlo errors were less than 10% of the standard error and FMI values were no larger than 0.7, suggesting that 100 imputed datasets were sufficient.^9^ Each imputation model contained all variables in the substantive analyses along with over 60 auxiliary variables related to socioeconomic adversity, parental characteristics (including maternal depression), and offspring psychopathology that have been identified as strong predictors of missingness in the outcome and confounders. We repeated our analyses by averaging parameter estimates across 100 imputed datasets and computing the associated standard error using Rubin’s rule.^10^ Our primary analyses examined the interaction between father absence and sex of the child, thus, it was necessary to incorporate the interaction term (father absence x sex) into the imputation model to ensure our estimates of the interaction effect are not attenuated. This was achieved by performing two separate imputations for each sex and combining the output prior to the analyses of the final model using the imputed datsets.^11^

For our trajectories analysis, we included individuals into our trajectories analyses if they had at least one measurement of depressive symptoms as previous research has shown that the shape and characteristics of the trajectories do not vary when including one measurement or at least four measurements of depressive symptoms.^12^ By default, missing outcome data (in this case depression) is handled using full information maximum likelihood (FIML), which assumes that the probability of an individual missing a measure of depressive symptoms does not depend on their underlying depressive symptoms score at that occasion, given their observed depressive symptoms trajectory at other occasions. Missing father absence data was handled using inverse-probability weighted analysis which weights each individual in the sample by the inverse probability of being sampled. We then used the Delta method (*nlcom* command) in Stata v.15/MP to estimate predicted scores between trajectories, with p-values for the predicted differences corrected using False Discovery Correction (FDR) to avoid bias from multiple comparisons.

**Results S1**

*Study sample derivation*

Of the 14,901 children who comprised the original ALSPAC sample, data on biological father presence versus absence from birth to age 10 years was available for 10,946. This comprised 8,277 (75.6%) children whose biological father was present from birth to age 10 years, 1,943 (17.7%) children whose father was absent from birth to 5 years, and 726 (6.6%) children whose father was absent from 5 to 10 years. Complete outcome data were available for 3,966 and 4,022 offspring with depression diagnosis and depressive symptoms at 24 years. Sample sizes with complete data on exposure, outcome and confounding factors to examine the effects of biological father absence on offspring depression diagnosis (father absence birth-5 years: n=2,672; father absence 5-10 years: n=2,435) and depressive symptoms (father absence birth-5 years: n=2,694; father absence 5-10 years: n=2,450) at age 24 years are presented in Figure 1. The sample sizes for those with data on exposure, confounding factors and at least one measurement of depressive symptoms (to establish trajectories) was 6,020 (father absence birth-5 years) and 5,352 (father absence 5-10 years).

*Characteristics of study sample by data completeness*

Characteristics of the sample by the completeness of the data are presented in Table S3. Sociodemographic differences between the samples indicated that our study sample comprised mothers who were from a more advantageous socioeconomic background, reporting higher levels of educational attainment, lower levels of depressive symptoms and parental conflict compared to mothers lost to follow-up.

*Distribution of depression, socioeconomic and familial characteristics in father-present and father-absent samples*

There were notable differences in the levels of socioeconomic resources and familial characteristics between father-present and father-absent families (Table S1). Mothers whose partners were absent reported lower levels of educational attainment, higher levels of depressive symptoms and inter-parental conflict than those whose partners were present. Father-absent families were also more likely to be of lower socioeconomic background, as indexed by the higher rate of manual social class, higher prevalence of financial difficulties and residence in council accommodation. Girls whose fathers were present between birth-5 years, but not absent, had a higher prevalence of depression diagnosis and depressive symptoms at age 24 years than boys (Table S2). Girls whose fathers were both present and absent between ages 5 and 10 years had higher prevalence of depression diagnosis and depressive symptoms than boys.

*Missing data: association between father absence in childhood and depression (diagnosis and symptoms) at age 24 years*

In order to examine the impact of response attrition on our findings, we imputed missing data in the outcome and confounders and repeated the analyses in the imputed samples. The results from the analyses with imputed data supported findings of our complete case analyses and led to the same over-arching conclusions. Specifically, there was stronger evidence of the main effect of father absence in early (OR: 2.16, 95%CI 1.65, 2.83, p≤0.001; Table S4), but not middle childhood (OR: 1.19, 95%CI 0.74, 1.95, p=0.464; Table S4), on depression diagnosis at age 24 years. Similarly to the complete case analyses, there was no evidence for an interaction between sex and father absence in early (birth-5 years) childhood (Table S4). There was also stronger evidence for a main effect of father absence in early (OR: 1.70, 95%CI 1.39, 2.10, p≤0.001; Table S4) and middle (OR: 1.40, 95% CI 1.06, 1.86, p=0.018; Table S4) childhood on depressive symptoms at age 24 years. There was no evidence for an interaction between sex and father absence in early (birth-5 years) or middle (5-10 years) childhood on depressive symptoms at age 24 years in imputed data analyses.

Consistent with the complete case analyses, there was strong evidence for the main effect of father absence in early childhood (birth-5 years) on depression diagnosis (OR: 1.67, 95%CI 1.25, 2.23, p=0.001; Table S5) and depressive symptoms (OR: 1.32, 95%CI 1.07, 1.64, p=0.010; Table S5) at age 24 years in the fully adjusted models. There was no evidence for the main effect of father absence in middle childhood (5-10 years) on depression diagnosis (OR: 1.04, 95%CI 0.65, 1.70, p=0.843; Table S5) or depressive symptoms (OR: 1.23, 95%CI 0.92, 1.64, p=0.165; Table S5) at age 24 years in the fully adjusted models.

**References**

1. Angold, A., Costello, E.J., Messer, S.C., Pickles, A., Winder, F., & Silver, D. (1995). Development of a short questionnaire for use in epidemiological studies of depression in children and adolescents. *International Journal of Methods in Psychiatric Research,* 5, 237–249.

2. Shaffer, D., Fisher, P., Lucas, C. P., Dulcan, M. K., & Schwab-Stone, M. E. (2000). NIMH Diagnostic Interview Schedule for Children Version IV (NIMH DISC-IV): description, differences from previous versions, and reliability of some common diagnoses. *Journal of the American Academy of Child & Adolescent Psychiatry*, *39*(1), 28-38.

3. Kwong, A. S., Morris, T. T., Pearson, R. M., Timpson, N. J., Rice, F., Stergiakouli, E., & Tilling, K. (2021). Polygenic risk for depression, anxiety and neuroticism are associated with the severity and rate of change in depressive symptoms across adolescence. *Journal of Child Psychology and Psychiatry*. Doi: https://doi.org/10.1111/jcpp.13422.

4. Kwong, A. S. (2019). Examining the longitudinal nature of depressive symptoms in the Avon Longitudinal Study of Parents and Children (ALSPAC) [version 2; peer review: 3 approved]. *Wellcome Open Research, 4*, 126. (<https://doi.org/10.12688/wellcomeopenres.15395.2>)

5. Stata v.15/MP. StataCorp., USA; [New in Stata 15 | Stata](https://www.stata.com/stata15/).
6. Leckie, G. and Charlton, C. (2013). [runmlwin - A Program to Run the MLwiN Multilevel Modelling Software from within Stata](http://www.jstatsoft.org/v52/i11). Journal of Statistical Software, 52 (11),1-40. ([do](http://www.bristol.ac.uk/cmm/media/runmlwin/jss.do)).
7. Sterne, J. A. C., White, I. R, Carlin, J. B, Spratt, M., Royston, P., Kenward, M. G., Wood, A. M., & Carpenter, J. R. (2009). Multiple imputation for missing data in epidemiological and clinical research: potential and pitfalls. *British Medical Journal, 338*, b2393-b2393.
8. White, I. R., Royston, P., & Wood, A. M. (2011). Multiple imputation using chained equations: issues and guidance for practice. *Statistics in medicine*, *30*(4), 377-399.
9. Rubin D. (1987). Multiple imputation for nonresponse in surveys. New York, USA: John Willey & Sons.
10. Van Buuren, S. (2012). Multiple imputation. In *Flexible Imputation of Missing Data,* Chapman and Hall/CRC.

11. Kwong, A. S., Manley, D., Timpson, N. J., Pearson, R. M., Heron, J., Sallis, H., Stergiakouli, E., Davis, O. S. P., & Leckie, G. (2019). Identifying critical points of trajectories of depressive symptoms from childhood to young adulthood. *Journal of Youth and Adolescence*, *48*(4), 815-827. <https://link.springer.com/article/10.1007/s10964-018-0976-5>.

**Table S1.** Characteristics of the father-absent (birth-5 and 5-10 years) and father-present samples

| Risk factors | | Father presence/absence  birth-5 years | | | Father presence/absence  5-10 years | | |
| --- | --- | --- | --- | --- | --- | --- | --- |
|  |  | Father present | Father absent |  | Father present | Father absent |  |
|  |  | n (%) | n (%) | p^a^ | n (%) | n (%) | p^a^ |
| Major financial problems | *No financial problems* | 6,547 (78.8) | 989 (59.7) | ≤0.001 | 6,104 (79.9) | 443 (66.4) | ≤0.001 |
|  | *Financial problems* | 1,761 (21.2) | 667 (40.3) |  | 1,537 (20.1) | 224 (33.6) |  |
| Maternal educational attainment | *University degree* | 3,506 (41.1) | 424 (24.7) | ≤0.001 | 3,271 (41.7) | 235 (34.6) | ≤0.001 |
|  | *A-Level* | 3,025 (35.5) | 623 (36.2) |  | 2,740 (34.9) | 285 (41.9) |  |
|  | *O-Level* | 1,993 (23.4) | 672 (39.1) |  | 1,833 (23.4) | 160 (23.5) |  |
| Parental social class | *Professional/managerial* | 2,978 (40.9) | 364 (28.1) | ≤0.001 | 2,757 (41.1) | 221 (38.8) | 0.289 |
|  | *Manual* | 4,296 (59.1) | 930 (71.9) |  | 3,948 (58.9) | 348 (61.2) |  |
| Homeownership status | *Owned/mortgaged* | 7,138 (85.5) | 890 (54.0) | ≤0.001 | 6,640 (86.4) | 498 (76.1) | ≤0.001 |
|  | *Private rented* | 416 (5.0) | 234 (14.2) |  | 367 (5.0) | 49 (7.5) |  |
|  | *Council rented* | 789 (9.5) | 523 (31.8) |  | 682 (8.9) | 107 (16.4) |  |
|  |  | Mean (SD) | Mean (SD) | P^b^ | Mean (SD) | Mean (SD) | P^b^ |
| Maternal antenatal depression (EPDS) | | 6.4 (4.5) | 8.55 (5.4) | ≤0.001 | 6.3 (4.5) | 7.5 (4.9) | ≤0.001 |
| Parental conflict | | 10.2 (1.7) | 9.31 (1.9) | ≤0.001 | 10.2 (1.7) | 9.7 (1.7) | ≤0.001 |

^a^P-value for Pearson’s Chi-square; ^b^p-value for two-sided T-test.

Note: Sample sizes vary because of differences in data availability on sociodemographic, parental and familial characteristics.

EPDS: Edinburgh Postnatal Depression Scale

**Table S2.** Distribution of depression (diagnosis^a^ and symptoms^b^) in the father-absent (birth-5 and 5-10 years) and father present samples by sex

| Risk factors | | Father presence/absence (birth-5 years) | | | | | | Father presence/absence (5-10 years) | | | | | |
| --- | --- | --- | --- | --- | --- | --- | --- | --- | --- | --- | --- | --- | --- |
|  |  | Father present | | | Father absent | | | Father present | | | Father absent | | |
|  |  | Boys  n (%) | Girls  n (%) | p^c^ | Boys  n (%) | Girls  n (%) | p^c^ | Boys  n (%) | Girls  n (%) | p^c^ | Boys  n (%) | Girls  n (%) | p^c^ |
| Depression diagnosis at 24 years | *No* | 1,141 (93.8) | 1,701 (88.0) | ≤0.001 | 125  (85.6) | 216  (80.6) | 0.200 | 1,068 (93.6) | 1,577 (88.2) | ≤0.001 | 70  (95.0) | 124  (85.5) | 0.017 |
|  | *Yes* | 76  (6.2) | 233  (12.0) |  | 21 (14.4) | 52 (19.4) |  | 73  (6.4) | 212  (11.8) |  | 6  (5.0) | 21  (14.5) |  |
| Depressive symptoms at 24 years | *No* | 953  (82.1) | 1,508  (73.7) | ≤0.001 | 92  (71.3) | 196  (66.0) | 0.281 | 894  (82.0) | 1,416  (74.5) | ≤0.001 | 59  (84.3) | 92  (63.0) | 0.001 |
|  | *Yes* | 208  (17.9) | 538  (26.3) |  | 37 (28.7) | 101  (34.0) |  | 197  (18.0) | 484  (25.5) |  | 11  (15.7) | 54  (37.0) |  |

^a^Assessed using Clinical Interview Schedule-Revised (CIS-R); ^b^assessed using Short Mood and Feeling Questionnaire (SMFQ) ≥10; ^c^ p-value for Pearson’s Chi-square.

**Table S3.** Distribution of sociodemographic characteristics in the original Avon Longitudinal Study of Parents and Children (ALSPAC) Cohort and the study complete and imputed samples

| Sample demographic characteristics measured during pregnancy^1^ | Core ALSPAC sample^a^ | Complete sample^b^ | | Imputed sample^c^ | |
| --- | --- | --- | --- | --- | --- |
|  |  | Father presence/  absence (birth-5 years) | Father presence/  absence (5-10 years) | Father presence/  absence (birth-5 years) | Father presence/  absence  (5-10 years) |
|  | (n=14,901) | (n=2,672) | (n=2,435) | (n=10,946) | (n=9,009) |
|  | (%) | (%) | (%) | (%) | (%) |
| *Major financial problems* |  |  |  |  |  |
| No financial problems | 74.0 | 83.0 | 84.3 | 75.0 | 78.5 |
| Financial problems | 26.0 | 17.0 | 15.7 | 25.0 | 21.5 |
| *Maternal educational attainment* |  |  |  |  |  |
| University degree | 35.3 | 52.6 | 53.8 | 38.0 | 40.8 |
| A-Level | 34.6 | 34.0 | 33.6 | 36.0 | 35.5 |
| O-Level | 30.1 | 3.4 | 12.6 | 26.0 | 23.7 |
| *Parental social class* |  |  |  |  |  |
| Professional/managerial | 37.4 | 52.7 | 48.7 | 36.1 | 38.5 |
| Manual | 62.6 | 47.3 | 51.3 | 63.9 | 61.5 |
| *Homeownership status* |  |  |  |  |  |
| Owned/mortgaged | 75.9 | 91.4 | 92.5 | 79.0 | 85.1 |
| Private rented | 7.5 | 4.0 | 3.6 | 7.0 | 5.1 |
| Council rented | 16.6 | 4.6 | 3.9 | 14.0 | 9.8 |
| *Maternal antenatal depression (EPDS)* |  |  |  |  |  |
| No (≤13) | 86.1 | 90.0 | 91.4 | 87.2 | 89.4 |
| Yes (≥13) | 13.9 | 10.0 | 8.6 | 12.8 | 10.6 |
| *Parental conflict* |  |  |  |  |  |
| No | 62.7 | 55.7 | 54.0 | 61.4 | 58.8 |
| Yes | 37.3 | 44.31 | 46.1 | 38.6 | 41.2 |

*Note*: ^1^Additional missing data on demographics: major financial problems missing for 2,748/(22.3%); maternal educational attainment missing for 2,418/(20.2%); parental social class missing for 4,792/(35.4%); homeownership status missing for 1,882/(16.8%); maternal antenatal depression missing for 2,569/(21.2%); parental conflict missing for 2,822/(22.8%).

^a^Core ALSPAC sample: no exposure or outcome data; ^b^Complete sample: exposure, outcomes and confounders data available; ^c^ Imputed sample: imputed missing data on outcome and confounders.

**Table S4.** Odds ratios for [95% Cis] for the association between father absence during different periods in childhood and binary indicators of depression diagnosis and depressive symptoms at 24 years stratifying by sex in the imputed samples

| Risk factor:  Timing of father absence | Depression diagnosis (CIS-R) at 24 years  (Reference=no depression diagnosis) | | |
| --- | --- | --- | --- |
|  | n | Main effect | Main effect+interaction term^a^ |
|  |  | OR [95% CI], p | OR [95% CI], p |
| Father left between birth-5 years  (Reference=Father present) | 10,946 | 2.16 [1.65, 2.83], p≤0.001 | 2.58 [1.58, 4.21], p≤0.001 |
| Gender (Reference=Male) |  | 1.63 [1.28, 2.07], p≤0.001 | 1.78 [1.39, 2.30], p≤0.001 |
| Father absence x sex |  | - | 0.73 [0.40, 1.29], p=0.271 |
| Father left between 5-10 years  (Reference=Father present) | 9,003 | 1.19 [0.74, 1.95], p=0.464 | 0.91 [0.30, 2.74], p=0.862 |
| Gender (Reference=Male) |  | 1.53 [1.12, 2.10], p=0.008 | 1.48 [1.08, 2.04], p=0.015 |
| Father absence x sex |  | - | 1.49 [0.44, 5.03], p=0.517 |
|  | Depressive symptoms (SMFQ) at 24 years  (Reference=no depressive symptoms) | | |
| Father left between birth-5 years  (Reference=Father present) | 10,946 | 1.70 [1.39, 2.10], p≤0.001 | 1.97 [1.35, 2.89], p=0.001 |
| Gender (Reference=Male) |  | 1.52 [1.26, 1.76], p≤0.001 | 1.58 [1.34, 1.87], p≤0.001 |
| Father absence x sex |  | - | 0.75 [0.46, 1.21], p=0.239 |
| Father left between 5-10 years  (Reference=Father present) | 9,003 | 1.40 [1.06, 1.86], p=0.018 | 1.06 [0.61, 1.84], p=0.824 |
| Gender (Reference=Male) |  | 1.55 [1.29, 1.85], p≤0.001 | 1.48 [1.23, 1.78], p≤0.001 |
| Father absence x sex |  | - | 1.61 [0.85, 3.04], p=0.140 |

^a^The interaction term: father absence by sex.

OR: odds ratio.

CIS-R: Clinical Interview Schedule-Revised.

SMFQ: Short Mood and Feelings Questionnaire.

**Table S5.** Odds ratios [95% CI] for the main effect of father absence during different periods in childhood on binary indicators of depression diagnosis and depressive symptoms at 24 years in the imputed sample

| Risk factor:  Timing of father absence | n | Depression diagnosis (CIS-R) at 24 years  (Reference=no depression diagnosis) | | Depressive symptoms (SMFQ) at 24 years  (Reference=no depressive symptoms) | |
| --- | --- | --- | --- | --- | --- |
| Father absence  (Reference=Father present) |  | Unadjusted  OR [95% CI], p | Adjusted^a^  OR [95% CI], p | Unadjusted  OR [95% CI], p | Adjusted^a^  OR [95% CI], p |
| Father left between birth-5 years | 10,946 | 2.16 [1.65, 2.83],  p≤0.001 | 1.67 [1.25, 2.23],  p=0.001 | 1.70 [1.39, 2.10],  p≤0.001 | 1.32 [1.07, 1.64],  p=0.010 |
| Father left between 5-10 years | 9,003 | 1.19 [0.74, 1.92],  p=0.464 | 1.04 [0.65, 1.70]  p=0.843 | 1.41 [1.10, 1.86],  p=0.018 | 1.23 [0.92, 1.64],  p=0.165 |

*Note*: ^a^Adjusted for antenatal indicators of socioeconomic (parental social class, financial difficulties, maternal educational attainment, homeownership), maternal (depression) and familial (parental conflict) characteristics.

OR: Odds ratio.

CIS-R: Clinical Interview Schedule-Revised.

SMFQ: Short Mood and Feelings Questionnaire.

**Table S6.** Estimates for the main effect of early father absence during childhood on trajectories of depressive symptoms (unadjusted)

|  | Estimates from unadjusted early father absence model (n=8,409) | | | | |
| --- | --- | --- | --- | --- | --- |
|  | Beta | Std Error | P-Value | Lower 95% CI | Upper 95% CI |
| No Father Absence Intercept | 6.120 | 0.064 | ≤0.001 | 5.996 | 6.245 |
| No Father Absence x Age | 0.350 | 0.015 | ≤0.001 | 0.320 | 0.380 |
| No Father Absence x Age^2^ | -0.096 | 0.005 | ≤0.001 | -0.106 | -0.087 |
| No Father Absence x Age^3^ | -0.005 | 0.0004 | ≤0.001 | -0.006 | -0.004 |
| No Father Absence x Age^4^ | 0.002 | 0.0001 | ≤0.001 | 0.002 | 0.002 |
| Early Father Absence Intercept | 0.997 | 0.178 | ≤0.001 | 0.649 | 1.346 |
| Early Father Absence x Age | 0.079 | 0.045 | 0.078 | -0.009 | 0.166 |
| Early Father Absence x Age^2^ | -0.021 | 0.014 | 0.137 | -0.049 | 0.007 |
| Early Father Absence x Age^3^ | -0.002 | 0.001 | 0.151 | -0.004 | 0.001 |
| Early Father Absence x Age^4^ | 0.0005 | 0.0003 | 0.078 | -0.0001 | 0.001 |

*Note:* Intercepts were set to age 16 to correspond with the average age of all assessments.

**Table S7.** Estimates for the main effect of early father absence during childhood on trajectories of depressive symptoms (adjusted)

|  | Estimates from adjusted early father absence model (n=6,020)^a^ | | | | |
| --- | --- | --- | --- | --- | --- |
|  | Beta | Std Error | P-Value | Lower 95% CI | Upper 95% CI |
| No Father Absence Intercept | 6.113 | 0.282 | ≤0.001 | 5.560 | 6.665 |
| No Father Absence x Age | 0.346 | 0.017 | ≤0.001 | 0.312 | 0.380 |
| No Father Absence x Age^2^ | -0.094 | 0.005 | ≤0.001 | -0.105 | -0.083 |
| No Father Absence x Age^3^ | -0.005 | 0.0004 | ≤0.001 | -0.006 | -0.004 |
| No Father Absence x Age^4^ | 0.002 | 0.0001 | ≤0.001 | 0.002 | 0.002 |
| Early Father Absence Intercept | 0.643 | 0.229 | 0.005 | 0.195 | 1.091 |
| Early Father Absence x Age | 0.067 | 0.057 | 0.240 | -0.045 | 0.179 |
| Early Father Absence x Age^2^ | -0.029 | 0.018 | 0.115 | -0.065 | 0.007 |
| Early Father Absence x Age^3^ | -0.001 | 0.001 | 0.411 | -0.004 | 0.002 |
| Early Father Absence x Age^4^ | 0.001 | 0.0004 | 0.071 | -0.0001 | 0.001 |

*Note:* Intercepts were set to age 16 to correspond with the average age of all assessments. ^a^Adjusted for antenatal indicators of socioeconomic (parental social class, financial difficulties, maternal educational attainment, homeownership), maternal (depression) and familial (parental conflict) characteristics.

**Table S8.** Estimates for the main effect of later father absence during childhood on trajectories of depressive symptoms (unadjusted)

|  | Estimates from unadjusted later father absence model (n=7,167) | | | | |
| --- | --- | --- | --- | --- | --- |
|  | Beta | Std Error | P Value | Lower 95% CI | Upper 95% CI |
| No Father Absence Intercept | 6.043 | 0.065 | ≤0.001 | 5.915 | 6.171 |
| No Father Absence x Age | 0.352 | 0.016 | ≤0.001 | 0.321 | 0.383 |
| No Father Absence x Age^2^ | -0.094 | 0.005 | ≤0.001 | -0.104 | -0.084 |
| No Father Absence x Age^3^ | -0.005 | 0.0004 | ≤0.001 | -0.006 | -0.004 |
| No Father Absence x Age^4^ | 0.002 | 0.0001 | ≤0.001 | 0.002 | 0.002 |
| Later Father Absence Intercept | 0.974 | 0.236 | ≤0.001 | 0.512 | 1.437 |
| Later Father Absence x Age | -0.024 | 0.059 | 0.684 | -0.139 | 0.091 |
| Later Father Absence x Age^2^ | -0.031 | 0.019 | 0.099 | -0.069 | 0.006 |
| Later Father Absence x Age^3^ | 0.001 | 0.001 | 0.414 | -0.002 | 0.004 |
| Later Father Absence x Age^4^ | 0.001 | 0.0004 | 0.156 | -0.0002 | 0.001 |

*Note:* Intercepts were set to age 16 to correspond with the average age of all assessments.

**Table S9.** Estimates for the main effect of later father absence during childhood on trajectories of depressive symptoms (adjusted)

|  | Estimates from adjusted later father absence model (n=5,352)^a^ | | | | |
| --- | --- | --- | --- | --- | --- |
|  | Beta | Std Error | P Value | Lower 95% CI | Upper 95% CI |
| No Father Absence Intercept | 5.965 | 0.298 | ≤0.001 | 5.382 | 6.549 |
| No Father Absence x Age | 0.347 | 0.018 | ≤0.001 | 0.312 | 0.382 |
| No Father Absence x Age^2^ | -0.092 | 0.006 | ≤0.001 | -0.103 | -0.081 |
| No Father Absence x Age^3^ | -0.005 | 0.0005 | ≤0.001 | -0.006 | -0.004 |
| No Father Absence x Age^4^ | 0.002 | 0.0001 | ≤0.001 | 0.001 | 0.002 |
| Later Father Absence Intercept | 0.682 | 0.273 | 0.012 | 0.148 | 1.216 |
| Later Father Absence x Age | -0.016 | 0.068 | 0.817 | -0.150 | 0.118 |
| Later Father Absence x Age^2^ | -0.030 | 0.022 | 0.178 | -0.073 | 0.013 |
| Later Father Absence x Age^3^ | 0.000 | 0.002 | 0.800 | -0.003 | 0.004 |
| Later Father Absence x Age^4^ | 0.001 | 0.0004 | 0.254 | -0.0004 | 0.001 |

*Note:* Intercepts were set to age 16 to correspond with the average age of all assessments. ^a^Adjusted for antenatal indicators of socioeconomic (parental social class, financial difficulties, maternal educational attainment, homeownership), maternal (depression) and familial (parental conflict) characteristics.

**Table S10.** Estimates for the main effect of early father absence during childhood on trajectories of depressive symptoms stratified by sex (unadjusted)

|  | Estimates from unadjusted early father absence by gender model (n=8,409) | | | | |
| --- | --- | --- | --- | --- | --- |
|  | Beta | Std Error | P Value | Lower 95% CI | Upper 95% CI |
| Male No Father Absence Intercept | 4.892 | 0.094 | ≤0.001 | 4.708 | 5.075 |
| Male No Father Absence x Age | 0.293 | 0.024 | ≤0.001 | 0.247 | 0.339 |
| Male No Father Absence x Age^2^ | -0.051 | 0.008 | ≤0.001 | -0.067 | -0.036 |
| Male No Father Absence x Age^3^ | -0.005 | 0.001 | ≤0.001 | -0.007 | -0.004 |
| Male No Father Absence x Age^4^ | 0.001 | 0.0002 | ≤0.001 | 0.001 | 0.002 |
| Female No Father Absence Intercept | 2.217 | 0.125 | ≤0.001 | 1.972 | 2.462 |
| Female No Father Absence x Age | 0.078 | 0.031 | 0.013 | 0.017 | 0.139 |
| Female No Father Absence x Age^2^ | -0.077 | 0.010 | ≤0.001 | -0.096 | -0.058 |
| Female No Father Absence x Age^3^ | 0.001 | 0.001 | 0.088 | -0.0002 | 0.003 |
| Female No Father Absence x Age^4^ | 0.001 | 0.0002 | ≤0.001 | 0.0004 | 0.001 |
| Male Early Father Absence Intercept | 0.657 | 0.269 | 0.014 | 0.130 | 1.184 |
| Male Early Father Absence x Age | -0.016 | 0.071 | 0.827 | -0.155 | 0.124 |
| Male Early Father Absence x Age^2^ | -0.025 | 0.024 | 0.286 | -0.072 | 0.021 |
| Male Early Father Absence x Age^3^ | 0.001 | 0.002 | 0.749 | -0.003 | 0.004 |
| Male Early Father Absence x Age^4^ | 0.001 | 0.0005 | 0.081 | -0.0001 | 0.002 |
| Female Early Father Absence Intercept | 3.381 | 0.232 | ≤0.001 | 2.927 | 3.835 |
| Female Early Father Absence x Age | 0.189 | 0.059 | 0.001 | 0.074 | 0.305 |
| Female Early Father Absence x Age^2^ | -0.098 | 0.018 | ≤0.001 | -0.134 | -0.062 |
| Female Early Father Absence x Age^3^ | -0.001 | 0.002 | 0.456 | -0.004 | 0.002 |
| Female Early Father Absence x Age^4^ | 0.001 | 0.0004 | 0.001 | 0.0004 | 0.002 |

*Note:* Intercepts were set to age 16 to correspond with the average age of all assessments.

**Table S11.** Estimates for the main effect of early father absence during childhood on trajectories of depressive symptoms stratified by sex (adjusted)

|  | Estimates from adjusted early father absence model (n=6,020)^a^ | | | | |
| --- | --- | --- | --- | --- | --- |
|  | Beta | Std Error | P Value | Lower 95% CI | Upper 95% CI |
| Male No Father Absence Intercept | 4.892 | 0.094 | ≤0.001 | 4.708 | 5.075 |
| Male No Father Absence x Age | 0.293 | 0.024 | ≤0.001 | 0.247 | 0.339 |
| Male No Father Absence x Age^2^ | -0.051 | 0.008 | ≤0.001 | -0.067 | -0.036 |
| Male No Father Absence x Age^3^ | -0.005 | 0.001 | ≤0.001 | -0.007 | -0.004 |
| Male No Father Absence x Age^4^ | 0.001 | 0.0002 | ≤0.001 | 0.001 | 0.002 |
| Female No Father Absence Intercept | 2.217 | 0.125 | ≤0.001 | 1.972 | 2.462 |
| Female No Father Absence x Age | 0.078 | 0.031 | 0.013 | 0.017 | 0.139 |
| Female No Father Absence x Age^2^ | -0.077 | 0.010 | ≤0.001 | -0.096 | -0.058 |
| Female No Father Absence x Age^3^ | 0.001 | 0.001 | 0.088 | -0.0002 | 0.003 |
| Female No Father Absence x Age^4^ | 0.001 | 0.0002 | ≤0.001 | 0.0004 | 0.001 |
| Male Early Father Absence Intercept | 0.657 | 0.269 | 0.014 | 0.130 | 1.184 |
| Male Early Father Absence x Age | -0.016 | 0.071 | 0.827 | -0.155 | 0.124 |
| Male Early Father Absence x Age^2^ | -0.025 | 0.024 | 0.286 | -0.072 | 0.021 |
| Male Early Father Absence x Age^3^ | 0.001 | 0.002 | 0.749 | -0.003 | 0.004 |
| Male Early Father Absence x Age^4^ | 0.001 | 0.0005 | 0.081 | -0.0001 | 0.002 |
| Female Early Father Absence Intercept | 3.381 | 0.232 | ≤0.001 | 2.927 | 3.835 |
| Female Early Father Absence x Age | 0.189 | 0.059 | 0.001 | 0.074 | 0.305 |
| Female Early Father Absence x Age^2^ | -0.098 | 0.018 | ≤0.001 | -0.134 | -0.062 |
| Female Early Father Absence x Age^3^ | -0.001 | 0.002 | 0.456 | -0.004 | 0.002 |
| Female Early Father Absence x Age^4^ | 0.001 | 0.0004 | 0.001 | 0.0004 | 0.002 |

*Note:* Intercepts were set to age 16 to correspond with the average age of all assessments. ^a^Adjusted for antenatal indicators of socioeconomic (parental social class, financial difficulties, maternal educational attainment, homeownership), maternal (depression) and familial (parental conflict) characteristics.

**Table S12.** Estimates for the main effect of later father absence during childhood on trajectories of depressive symptoms stratified by sex (unadjusted)

|  | Estimates from unadjusted later father absence model (n=7,167) | | | | |
| --- | --- | --- | --- | --- | --- |
|  | Beta | Std Error | P Value | Lower 95% CI | Upper 95% CI |
| Male No Father Absence Intercept | 4.881 | 0.096 | ≤0.001 | 4.693 | 5.068 |
| Male No Father Absence x Age | 0.304 | 0.024 | ≤0.001 | 0.257 | 0.351 |
| Male No Father Absence x Age^2^ | -0.051 | 0.008 | ≤0.001 | -0.067 | -0.036 |
| Male No Father Absence x Age^3^ | -0.006 | 0.001 | ≤0.001 | -0.007 | -0.004 |
| Male No Father Absence x Age^4^ | 0.001 | 0.0002 | ≤0.001 | 0.001 | 0.002 |
| Female No Father Absence Intercept | 2.112 | 0.128 | ≤0.001 | 1.860 | 2.363 |
| Female No Father Absence x Age | 0.065 | 0.032 | 0.043 | 0.002 | 0.128 |
| Female No Father Absence x Age^2^ | -0.074 | 0.010 | ≤0.001 | -0.094 | -0.054 |
| Female No Father Absence x Age^3^ | 0.001 | 0.001 | 0.075 | -0.0001 | 0.003 |
| Female No Father Absence x Age^4^ | 0.001 | 0.0002 | ≤0.001 | 0.0004 | 0.001 |
| Male Later Father Absence Intercept | 0.099 | 0.366 | 0.787 | -0.619 | 0.817 |
| Male Later Father Absence x Age | -0.167 | 0.095 | 0.078 | -0.353 | 0.019 |
| Male Later Father Absence x Age^2^ | -0.008 | 0.033 | 0.813 | -0.072 | 0.056 |
| Male Later Father Absence x Age^3^ | 0.003 | 0.002 | 0.213 | -0.002 | 0.008 |
| Male Later Father Absence x Age^4^ | 0.0003 | 0.001 | 0.634 | -0.001 | 0.002 |
| Female Later Father Absence Intercept | 3.521 | 0.300 | ≤0.001 | 2.932 | 4.110 |
| Female Later Father Absence x Age | 0.089 | 0.077 | 0.243 | -0.061 | 0.240 |
| Female Later Father Absence x Age^2^ | -0.113 | 0.024 | ≤0.001 | -0.160 | -0.067 |
| Female Later Father Absence x Age^3^ | 0.003 | 0.002 | 0.175 | -0.001 | 0.007 |
| Female Later Father Absence x Age^4^ | 0.001 | 0.0005 | 0.007 | 0.0004 | 0.002 |

*Note:* Intercepts were set to age 16 to correspond with the average age of all assessments.

**Table S13.** Estimates for the main effect of later father absence during childhood on trajectories of depressive symptoms stratified by sex (adjusted)

|  | Estimates from adjusted later father absence by gender model (n=5,352)^a^ | | | | |
| --- | --- | --- | --- | --- | --- |
|  | Beta | Std Error | P Value | Lower 95% CI | Upper 95% CI |
| Male No Father Absence Intercept | 4.892 | 0.094 | ≤0.001 | 4.708 | 5.075 |
| Male No Father Absence x Age | 0.293 | 0.024 | ≤0.001 | 0.247 | 0.339 |
| Male No Father Absence x Age^2^ | -0.051 | 0.008 | ≤0.001 | -0.067 | -0.036 |
| Male No Father Absence x Age^3^ | -0.005 | 0.001 | ≤0.001 | -0.007 | -0.004 |
| Male No Father Absence x Age^4^ | 0.001 | 0.0002 | ≤0.001 | 0.001 | 0.002 |
| Female No Father Absence Intercept | 2.217 | 0.125 | ≤0.001 | 1.972 | 2.462 |
| Female No Father Absence x Age | 0.078 | 0.031 | 0.013 | 0.017 | 0.139 |
| Female No Father Absence x Age^2^ | -0.077 | 0.010 | ≤0.001 | -0.096 | -0.058 |
| Female No Father Absence x Age^3^ | 0.001 | 0.001 | 0.088 | -0.0002 | 0.003 |
| Female No Father Absence x Age^4^ | 0.001 | 0.0002 | ≤0.001 | 0.0004 | 0.001 |
| Male Later Father Absence Intercept | 0.657 | 0.269 | 0.014 | 0.130 | 1.184 |
| Male Later Father Absence x Age | -0.016 | 0.071 | 0.827 | -0.155 | 0.124 |
| Male Later Father Absence x Age^2^ | -0.025 | 0.024 | 0.286 | -0.072 | 0.021 |
| Male Later Father Absence x Age^3^ | 0.001 | 0.002 | 0.749 | -0.003 | 0.004 |
| Male Later Father Absence x Age^4^ | 0.001 | 0.0005 | 0.081 | -0.0001 | 0.002 |
| Female Later Father Absence Intercept | 3.381 | 0.232 | ≤0.001 | 2.927 | 3.835 |
| Female Later Father Absence x Age | 0.189 | 0.059 | 0.001 | 0.074 | 0.305 |
| Female Later Father Absence x Age^2^ | -0.098 | 0.018 | ≤0.001 | -0.134 | -0.062 |
| Female Later Father Absence x Age^3^ | -0.001 | 0.002 | 0.456 | -0.004 | 0.002 |
| Female Later Father Absence x Age^4^ | 0.001 | 0.0004 | 0.001 | 0.0004 | 0.002 |

*Note:* Intercepts were set to age 16 to correspond with the average age of all assessments. ^a^Adjusted for antenatal indicators of socioeconomic (parental social class, financial difficulties, maternal educational attainment, homeownership), maternal (depression) and familial (parental conflict) characteristics.

**Table S14.** Predicted mean depressive symptoms scores [95% CI] at various ages for the main effect of father absence during different periods in childhood on trajectories of depressive symptoms across all models

|  | Predicted Mean Depressive Symptoms Scores (SMFQ) at Various Ages^a^ | | | |
| --- | --- | --- | --- | --- |
| *Model 1* | Age 12 | Age 16 | Age 20 | Age 24 |
| No Father Absence (n=5,352) | 3.81 (3.26, 4.35) | 5.89 (5.34, 6.44) | 6.23 (5.67, 6.78) | 6.66 (6.09, 7.24) |
| Early Father Absence (n=668) | 3.94 (3.34, 4.54) | 6.48 (5.83, 7.14) | 6.80 (6.07, 7.53) | 7.72 (6.89, 8.55) |
| *Model 2* |  |  |  |  |
| No Father Absence (n=4,932) | 3.68 (3.10, 4.24) | 5.72 (5.14, 6.31) | 6.11 (5.52, 6.70) | 6.49 (5.88, 7.09) |
| Later Father Absence (n=420) | 3.99 (3.32, 4.65) | 6.41 (5.67, 7.14) | 6.48 (5.65, 7.31) | 7.12 (6.13, 8.11) |
| *Model 3* |  |  |  |  |
| Male No Early Father Absence (n=2,643) | 3.61 (3.06, 4.16) | 4.76 (4.19, 5.32) | 5.35 (4.76, 5.94) | 5.90 (5.27, 6.53) |
| Female No Early Father Absence (n=2,709) | 4.01 (3.46, 4.52) | 6.82 (6.26, 7.38) | 6.92 (6.34, 7.49) | 7.26 (6.67, 7.86) |
| Male Yes Early Father Absence (n=318) | 3.60 (2.92, 4.28) | 5.28 (4.49, 6.07) | 5.60 (4.63, 6.57) | 8.01 (6.78, 9.23) |
| Female Yes Early Father Absence (n=350) | 4.26 (3.61, 4.92) | 7.44 (6.71, 8.17) | 7.60 (6.78, 8.42) | 7.77 (6.81, 8.72) |
| *Model 4* |  |  |  |  |
| Male No Later Father Absence (n=2,441) | 3.50 (2.92, 4.08) | 4.69 (4.10, 5.29) | 5.31 (4.69, 5.92) | 5.82 (5.16, 6.48) |
| Female No Later Father Absence (n=2,491) | 3.89 (3.31, 4.47) | 6.63 (6.04, 7.22) | 6.76 (6.15, 7.36) | 7.10 (6.47, 7.73) |
| Male Yes Later Father Absence (n=202) | 3.79 (3.02, 4.56) | 4.35 (3.42, 5.29) | 4.64 (3.48, 5.79) | 5.76 (4.21, 7.31) |
| Female Yes Later Father Absence (n=218) | 4.30 (3.56, 5.05) | 7.89 (7.06, 8.72) | 7.68 (6.73, 8.62) | 8.16 (7.00, 9.32) |

*Note*: ^a^Adjusted for antenatal indicators of socioeconomic (parental social class, financial difficulties, maternal educational attainment, homeownership), maternal (depression) and familial (parental conflict) characteristics.

SMFQ: Short Mood and Feelings Questionnaire. **Table S15.** Inverse probability weighted (IPW) analysis estimates for the main effect of early father absence during childhood on trajectories of depressive symptoms (adjusted)

|  | IPW estimates from adjusted early father absence model (n=6,020)^a^ | | | | |
| --- | --- | --- | --- | --- | --- |
|  | Beta | Std Error | P Value | Lower 95% CI | Upper 95% CI |
| No Father Absence Intercept | 6.119 | 0.293 | ≤0.001 | 5.545 | 6.693 |
| No Father Absence x Age | 0.348 | 0.017 | ≤0.001 | 0.314 | 0.382 |
| No Father Absence x Age^2^ | -0.095 | 0.005 | ≤0.001 | -0.105 | -0.084 |
| No Father Absence x Age^3^ | -0.005 | 0.0004 | ≤0.001 | -0.006 | -0.004 |
| No Father Absence x Age^4^ | 0.002 | 0.0001 | ≤0.001 | 0.002 | 0.002 |
| Early Father Absence Intercept | 0.636 | 0.247 | 0.010 | 0.153 | 1.120 |
| Early Father Absence x Age | 0.068 | 0.061 | 0.265 | -0.051 | 0.187 |
| Early Father Absence x Age^2^ | -0.029 | 0.021 | 0.159 | -0.069 | 0.011 |
| Early Father Absence x Age^3^ | -0.001 | 0.002 | 0.434 | -0.004 | 0.002 |
| Early Father Absence x Age^4^ | 0.001 | 0.0004 | 0.103 | -0.0001 | 0.001 |

*Note:* Intercepts were set to age 16 to correspond with the average age of all assessments. ^a^Adjusted for antenatal indicators of socioeconomic (parental social class, financial difficulties, maternal educational attainment, homeownership), maternal (depression) and familial (parental conflict) characteristics.

**Table S16.** Inverse probability weighted (IPW) analysis estimates for the main effect of later father absence during childhood on trajectories of depressive symptoms (adjusted)

|  | IPW estimates from adjusted later father absence model (n=5,352)^a^ | | | | |
| --- | --- | --- | --- | --- | --- |
|  | Beta | Std Error | P Value | Lower 95% CI | Upper 95% CI |
| No Father Absence Intercept | 6.064 | 0.318 | ≤0.001 | 5.440 | 6.688 |
| No Father Absence x Age | 0.359 | 0.018 | ≤0.001 | 0.323 | 0.395 |
| No Father Absence x Age^2^ | -0.094 | 0.006 | ≤0.001 | -0.105 | -0.083 |
| No Father Absence x Age^3^ | -0.005 | 0.0005 | ≤0.001 | -0.006 | -0.004 |
| No Father Absence x Age^4^ | 0.002 | 0.0001 | ≤0.001 | 0.002 | 0.002 |
| Later Father Absence Intercept | 0.614 | 0.308 | 0.046 | 0.011 | 1.217 |
| Later Father Absence x Age | -0.031 | 0.077 | 0.691 | -0.182 | 0.120 |
| Later Father Absence x Age^2^ | -0.027 | 0.022 | 0.221 | -0.072 | 0.017 |
| Later Father Absence x Age^3^ | 0.001 | 0.002 | 0.736 | -0.003 | 0.005 |
| Later Father Absence x Age^4^ | 0.0004 | 0.0005 | 0.321 | -0.0004 | 0.001 |

*Note:* Intercepts were set to age 16 to correspond with the average age of all assessments. ^a^Adjusted for antenatal indicators of socioeconomic (parental social class, financial difficulties, maternal educational attainment, homeownership), maternal (depression) and familial (parental conflict) characteristics.
